# Human papillomavirus vaccine acceptability in Cameroon: a systematic review and meta-analysis

**DOI:** 10.64898/2026.04.28.26351975

**Authors:** Fabrice Zobel Lekeumo Cheuyem, Chabeja Achangwa, Rick Tchamani, Armand Tiotsia Tsapi, Edwige Omona Guissana, Eric Mboke Ekoum, Isabelle Nkwele Mekone

## Abstract

**Background:** Human papillomavirus (HPV) vaccination is a key strategy for cervical cancer elimination. In Cameroon, HPV vaccine was introduced into the expanded program on immunization in 2020. However, synthesized evidence on vaccine acceptability is needed to guide policy. This systematic review and meta-analysis aimed to estimate the pooled prevalence of HPV vaccine awareness, willingness to vaccinate, recommendation practices, and actual uptake in Cameroon, and to identify determinants of vaccine hesitancy.

**Methods:** We searched PubMed, Scopus, Web of Science, Embase, Cochrane Library, and African Journals Online from studies to January 2025. Studies reporting willingness to vaccinate, awareness, recommendation, and uptake of HPV vaccine were included. Pooled prevalence estimates and odds ratios were calculated using random-effects models. Heterogeneity was assessed using the *I*² statistic. The study was reported following PRISMA 2020 guidelines and registered in PROSPERO ID: CRD420261301213.

**Results:** Thirty-three studies were included. The pooled prevalence of willingness to vaccinate was 68.1% (95% CI: 57.4-77.2; 12 studies; n = 4,993; *I*² = 98%), while HPV vaccine awareness was 41.3% (95% CI: 28.7-55.1; 33 studies; n = 8,175 participants; *I*² = 98%). Two-thirds of participants (67.7%; 95% CI: 50.7-81.0; 8 studies; n = 1,617) reported recommending the vaccine, but actual uptake was only 22.9% (95% CI: 6.9-54.5; 9 studies; n = 9,686). Willingness significantly declined from 74.2% before 2014 to 57.5% after 2021. Healthcare workers had the highest awareness (74.5%) and willingness (77.8%). Lack of HPV knowledge was associated with nearly three-fold higher hesitancy (OR: 2.58; 95% CI: 2.06-3.22).

**Conclusions:** Despite moderate willingness, HPV vaccine awareness and uptake remain low in Cameroon, with marked disparities across regions and populations. Addressing knowledge gaps and strengthening context-specific vaccination strategies are needed to improve coverage.

## 1. Background

Human papillomavirus (HPV) is the most common sexually transmitted infection worldwide, with almost all sexually active individuals likely to contract it at some point of their life [1]. It is the main cause of cervical cancer globally. Cervical cancer is still a major public health issue globally, with about 90% of deaths happening in low- and middle-income countries [2]. Sub-Saharan Africa faces a higher burden of Cervical cancer because of limited screening, late diagnoses, and poor access to preventive care [3]. In Cameroon, cervical cancer is the second most common cancer among women and a leading cause of cancer-related deaths. Although highly preventable, the disease causes an estimated 1,700 to 1,800 deaths annually [4].

HPV vaccination is a very effective way to prevent cervical cancer. In 2020, the World Health Organization (WHO) introduced the Global Strategy to Accelerate the Elimination of Cervical Cancer, which set the “90–70–90” goals. One of these goals is to reach 90% HPV vaccination coverage among girls by age 15 by 2030 [5]. Meeting this goal needs more than just having the vaccine available; it also depends on public trust, acceptance, and good integration into national immunization programs.

HPV vaccination began in Cameroon around 2014, with pilot and demonstration projects in certain regions [6]. These early projects showed that awareness and acceptance were relatively high, especially among adolescents and parents who participated in organized information campaigns [7, 8]. However, these projects often received additional support and resources, which may not be the case during the regular national rollout.

After pilot studies in various regions of Cameroon in 2014, the vaccine was finally introduced into the national expanded program of immunization in 2020, with the main objectives of improving access and making the program more sustainable [9]. However, recent evidence shows that vaccination rates are still below WHO targets. While many studies report moderate to high willingness to vaccinate and recommend the vaccine, actual uptake is much lower in many communities [10, 11]. This gap between what people say and what happens in practice points to barriers, lack of information, and vaccine hesitancy.

Research from across Cameroon shows that awareness and acceptance of the HPV vaccine vary widely. For instance, healthcare workers and students usually have higher awareness and more positive attitudes, while women in the general population and some communities show lower awareness and more hesitancy [12, 13]. Not knowing about HPV infection and cervical cancer is a key reason for vaccine hesitancy in Cameroon [13]. This shows how health education and clear communication are important for improving vaccine uptake after integration to expanded program of immunization.

Even after intensive research on HPV vaccine awareness, willingness, recommendations, and uptake in Cameroon, the results are still mixed and vary by region, time, and group. There is also little research that looks at trends across the main phases: before vaccination (<2014), the demonstration phase (2014–2020), and after the vaccine was added to the EPI in 2020. A thorough review is needed to measure national acceptance and coverage and to guide strategies for improving HPV vaccination in the regular immunization system. This systematic review and meta-analysis aimed to estimate the overall rates of HPV vaccine awareness, willingness to vaccinate, recommendation practices, and actual uptake in Cameroon, and to identify the main factors associated with HPV vaccine hesitancy.

## 2. Methods

### 2.1. Study design and protocol registration

The systematic review and meta-analysis were reported based on the Preferred Reporting Items for Systematic Reviews and Meta-analysis (PRISMA) 2020. The study was registered with the international systematic review registry, PROSPERO ID CRD420261301213.

### 2.2. Eligibility Criteria

#### Inclusion criteria

This review focuses on quantitative, observational studies conducted before 2025. Interventional studies that measure at least our outcome of interest were also eligible. To be included, research must have reported on at least one of the following outcomes: HPV vaccine awareness, willingness to vaccinate against HPV, HPV vaccine recommendation, or HPV vaccine uptake among individuals residing in Cameroon. Finally, the review was limited to literature written in either English or French.

#### Exclusion criteria

Studies conducted outside Cameroon, qualitative studies, review articles, editorials, commentaries, conference abstracts, and letter to editor were excluded. Studies lacking sufficient data (full-text) to estimate these specific prevalences were also disregarded.

## 3. Search strategy

### 3.1. Databases

A comprehensive literature search was conducted in PubMed, Scopus, Cochrane library, Web of Science, Embase, and African Journals Online (AJOL), Health Sciences and Diseases. Additional studies were identified through Google Scholar and screening of reference lists of included articles. Screening was conducted by two study investigators (FZLC and RT) and disagreement solved through discussion.

### 3.2. Search terms

The search strategy used Medical Subject Headings (MeSH) and free-text terms combined with Boolean operators: (“human papillomavirus” OR HPV OR “cervical cancer”) AND (vaccine OR vaccination OR knowledge OR awareness OR attitude OR practice) AND (Cameroon OR Cameroonian OR Cameroun). For the PubMed repository the searching strategy was as follow: (("human papillomavirus"[TIAB] OR "HPV"[TIAB], "cervical cancer"[TIAB])) AND ("vaccine"[TIAB] OR "vaccination"[TIAB] OR "vaccines"[TIAB] OR "knowledge"[TIAB] OR "awareness"[TIAB] OR "attitude"[TIAB] OR "practice"[TIAB]) AND ("Cameroon"[Mesh] OR "Cameroon"[TIAB] OR "Cameroonian"[TIAB] OR "Cameroun"[TIAB])). The last search was conducted on January 27, 2025 (**Supplementary Table 1**).

### 3.3. Data extraction

Data was extracted using a standardized data extraction form. Extracted variables will include study characteristics (author, year of study completion, region, setting, and design), population characteristics, sampling methods, sample size, and reported outcomes of interest. Odds ratio (OR) and corresponding confidence intervals (CI) of factors associated with willingness to vaccinate was also extracted. Outcomes of interest included willingness or trust to vaccinate against HPV, knowledge of the HPV vaccine, HPV vaccine recommendation, HPV vaccine uptake. Data were extracted by two study investigators (FZLC, RT). and any disagreement solved through discussions or by consulting a third reviewer (CA).

### 3.4. Outcome measurement and operational definition

Our primary outcome of interest included the prevalence of willingness or trust to vaccinate against HPV which refer to individual who accepted to receive the HPV vaccine, or to have their children vaccinated against the HPV. Our secondary outcomes encompassed prevalence of knowledge of the HPV vaccine that prevent HPV infection and ultimately the cervical cancer; the prevalence of HPV vaccine recommendation which referred to the proportion of individuals who reported they can recommend HPV vaccine to other members of the community; and the prevalence of HPV vaccine uptake which represented the proportion of eligible individuals who reported ever received the HPV vaccine.

### 3.5. Data quality assessment

The risk of bias in the included articles will be assessed using the Joana Briggs Institute critical appraisal checklist [14]. The number of parameters used to assess the quality of the included manuscript will depend on the study design. For studies reporting prevalence data the items included : (1) sample frame appropriate to address the target population, (2) study participants sampled in an appropriate way, (3) sample size adequate, (4) study subjects and the setting described in detail, (5) data analysis conducted with sufficient coverage of the identified sample, (6) valid methods used for the identification of the condition, (7) condition measured in a standard, reliable way for all participants, (8) appropriate statistical analysis, (9) response rate adequate, and or low response rate managed appropriately. Each parameter will be scored as 1 (yes) or 0 (no or unclear). The risk of bias will be categorized as low: 7-9, moderate: 6-5, or high: 3-0 [15, 16]. Two reviewers independently assess the risk of bias. Disagreement between the reviewers were resolved by discussion to reach consensus (FZLC, ATT).

### 3.6. Statistical analysis and synthesis

For each study outcome, prevalence estimates and corresponding 95% confidence intervals (CIs) were calculated. To explore the variability in results, subgroup analyses were performed based on study timeframe, population group, study setting, sampling method. The *I^2^* statistic was used to assess heterogeneity between studies. A Der Simonian and Laird random-effects model was chosen to pool the estimates, anticipating significant heterogeneity. The Generalized Linear Mixed Models (GLMM), coupled with the Probit-Logit Transformation (PLOGIT), were utilized for their effectiveness in handling meta-analyses of binary data [17]. Statistical significance was set at a *p*-value of <0.05. All analyses were conducted using the ‘meta’ package in R Statistics version 4.5.2 [18]. Correlates to willingness to vaccinate against HPV were also computed to generate pooled ORs and their corresponding 95% CIs.

### 3.7. Publication bias and sensitivity analysis

Publication bias was assessed using funnel plots and statistically through Egger’s or Begg’s test, provided at least 10 studies were included in the meta-analysis [19, 20]. A *p*-value > 0.05 was considered indicative of no statistically significant evidence of publication bias. To evaluate the robustness of pooled estimates, a sensitivity analysis was performed using leave-one-out method (iteratively excluding one study at a time). The Duval and Tweedie trim-and-fill method was used to adjust for potential publication bias [21].

## 3. Results

### 3.1. Studies selection

A total of 448 articles were identified through searches of relevant databases. Of these, 25 duplicates were removed. After screening titles and abstracts, 346 reports were excluded. Among the remaining 77 full-text articles, 44 were excluded with specific reasons. Finally, 33 studies met the inclusion criteria and were included in the systematic review and meta-analysis (**Fig. 1**).

**Fig. 1.**
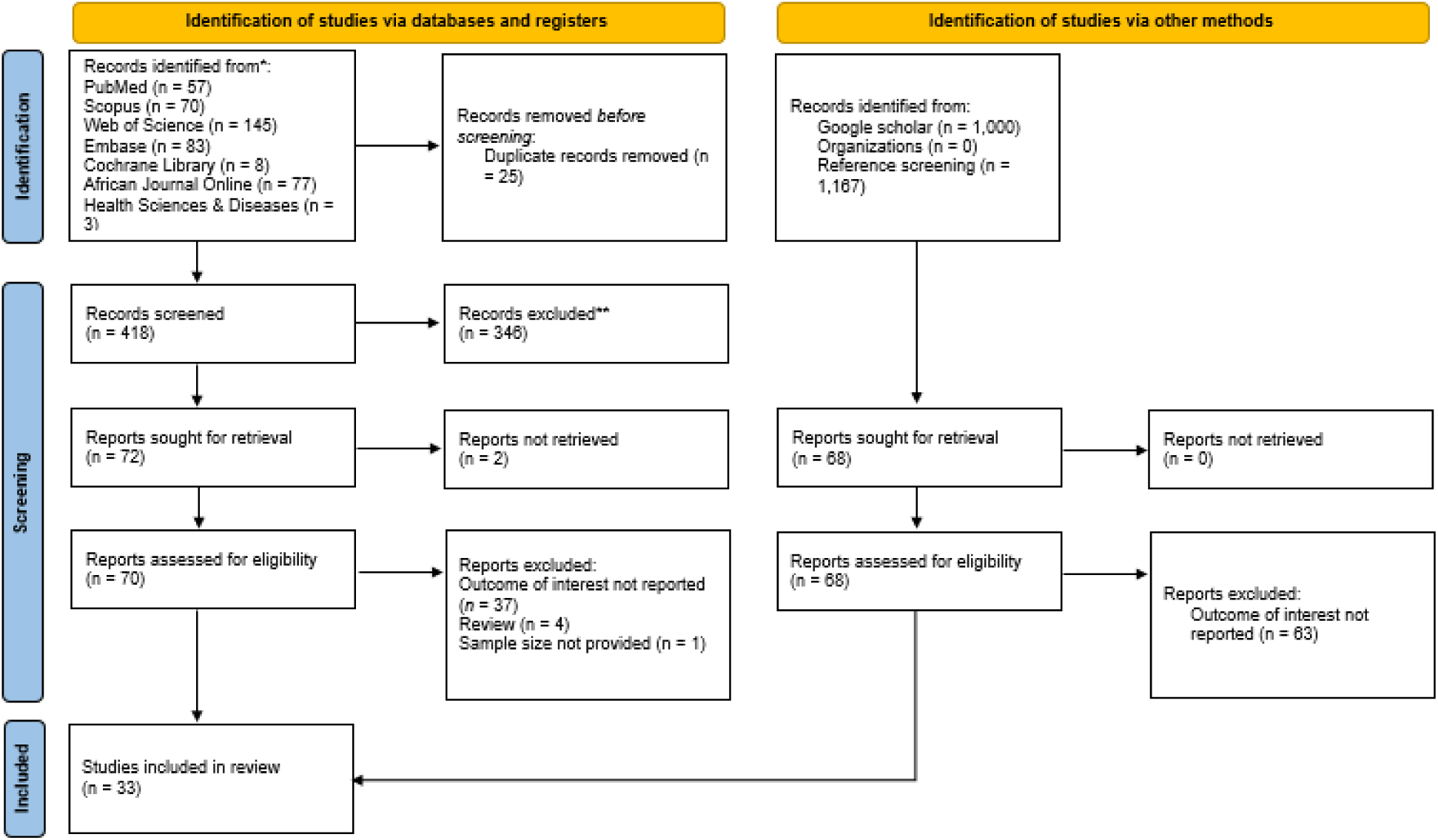
PRISMA diagram flow from study identification to inclusion in the systematic review and meta-analysis [22].

### 3.2. Study characteristics

A total of 33 study reports were included in this systematic review and meta-analysis. Most included studies were cross-sectional (n = 32; 97%) and were conducted from 2014 to 2020, corresponding to the HPV vaccination demonstration period in Cameroon (n = 21; 64%). The most frequently studied Cameroonian regions included the Centre (n = 16; 48%), followed by the West (n = 8; 24%), the South-West (n = 5; 15%) regions. Included reports were primarily hospital-based (n = 25; 76%) and used a non-probabilistic sampling method (n = 31; 94%). Most study participants were women belonging to an unspecified risk group or the general population (n = 25; 76%) (**Table 1**).

**Table 1.** Synthesis of included study characteristics.

| Author | Year<sup>1</sup> | Design | Region | Setting | Sampling method | Type of participant | Risk of bias | Sample size | Prevalence of HPV-related events (%) |  |  |  |
| --- | --- | --- | --- | --- | --- | --- | --- | --- | --- | --- | --- | --- |
|  |  |  |  |  |  |  |  |  | Knowledge of vaccine<sup>2</sup> | Willingness to vaccinate<sup>3</sup> | Recommend vaccine<sup>4</sup> | Vaccine uptake |
| McCarey,2009 [12] | 2009 | CS | Centre | Hospital | Non-probabilistic | Healthcare worker | Moderate | 401 | 44.14 | 65.59 | 88.70 | 60.35 |
| Ayissi,2011 [7] | 2011 | CS | North-West | School | Non-probabilistic | Adolescent girl | Moderate | 553 | 75.95 | 78.84 | 73.96 | 34.18 |
| Ladner,2011 [23] | 2011 | CS | NR | School and hospital | Non-probabilistic | Adolescent girl | Low | 6400 | NR | NR | NR | 90.56 |
| Wamai,2011_1 [24] | 2011 | CS | Centre & North-West | Hospital | Non-probabilistic | Healthcare worker | Moderate | 76 | 78.95 | NR | 69.74 | NR |
| Wamai,2011_2 [8] | 2011 | CS | North-West | Community | Non-probabilistic | Parent of eligible children | Low | 337 | 80.12 | 76.85 | 78.93 | NR |
| Sossauer,2012 [25] | 2012 | RCT | Centre | Community | Probabilistic | Women | Moderate | 301 | 57.81 | NR | NR | NR |
| Hallen-Ekane,2017 [26] | 2017 | CS | South-West | Hospital | Probabilistic | Student (male and female) | Low | 416 | 24.28 | NR | NR | NR |
| Mforteh,2017 [27] | 2017 | CS | North-West | Community | Non-probabilistic | Women | Low | 476 | 7.14 | NR | NR | NR |
| Simo,2017 [28] | 2017 | CS | West | Hospital | Probabilistic | Women | Moderate | 228 | 2.63 | NR | NR | NR |
| Kindong,2018 [29] | 2018 | CS | Littoral | Community | Probabilistic | Women | Low | 226 | 4.42 | NR | NR | NR |
| Layu,2018 [30] | 2018 | CS | North-West | Community | Probabilistic | Women | Low | 253 | 15.81 | NR | NR | NR |
| Nkfusai,2018 [31] | 2018 | CS | South-West | Community | Non-probabilistic | Women | Low | 434 | 53.92 | NR | NR | NR |
| Dekenyo,2018 [32] | 2018 | CS | West | Community | Probabilistic | Women at reproductive age | Low | 236 | 7.63 | NR | NR | NR |
| Eakin,2018 [33] | 2018 | CS | South-West | University | Non-probabilistic | Student female | Low | 120 | 66.67 | NR | NR | 1.67 |
| Fru,2019 [34] | 2019 | CS | South-West | Community | Non-probabilistic | Women | Low | 117 | 75.21 | NR | NR | NR |
| Guenou,2019 [10] | 2019 | CS | Centre | Hospital | Non-probabilistic | Parent of eligible children | Moderate | 168 | 14.88 | 80.36 | 58.33 | NR |
| Maïdadi,2019 [35] | 2019 | CS | Centre | Community | Non-probabilistic | Women at reproductive age | Low | 204 | 19.12 | NR | NR | NR |
| Guenou,2019 [10] | 2020 | CS | Centre | Hospital | Non-probabilistic | Healthcare worker | Moderate | 89 | 44.94 | 84.27 | NR | NR |
| Haddison,2020 [36] | 2020 | CS | Centre | Hospital | Non-probabilistic | Healthcare worker | Moderate | 24 | 91.67 | 75.00 | 75.00 | NR |
| Ambori,2020 [37] | 2020 | CS | South-West | Hospital | Non-probabilistic | Women | Low | 238 | 39.92 | NR | NR | NR |
| Sobe,2022 [38] | 2021 | CS | South-West | University | Non-probabilistic | Student (male and female) | Low | 414 | 56.28 | NR | NR | NR |
| Tassang,2021_1 [39] | 2021 | CS | Far North | Community | Non-probabilistic | Women | Low | 251 | 76.10 | NR | NR | NR |
| Tassang,2021_2 [40] | 2021 | CS | South-West | Community | Non-probabilistic | Women | Low | 250 | 40.40 | 40.40 | 17.20 | NR |
| Fru,2021 [11] | 2021 | CS | South-West | Community | Probabilistic | General population | Low | 250 | NR | NR | NR | 1.60 |
| Eta,2021 [41] | 2021 | CS | Littoral | Community | Non-probabilistic | Female sex worker | Low | 384 | 38.02 | NR | NR | NR |
| Tchouaket,2021 [42] | 2021 | CS | Centre and Littoral | Hospital | Non-probabilistic | Non-pregnant women | Low | 63 | NR | NR | NR | 69.84 |
| Afegenwi,2022 [43] | 2022 | CS | Multi-region | University | Probabilistic | Student (male and female) | Low | 1263 | 38.16 | NR | NR | 15.20 |
| Batoum,2023 [44] | 2023 | CS | Centre | Hospital | Non-probabilistic | Healthcare worker | Low | 28 | 92.86 | NR | NR | NR |
| Zeuko'o,2023 [45] | 2023 | CS | South-West | University | Probabilistic | Female student | Low | 500 | 45.40 | NR | NR | NR |
| Nechi,2023 [46] | 2023 | CS | Centre | Community | Probabilistic | Parent of eligible children | Low | 330 | NR | 26.97 | NR | NR |
| Nechi,2023 [46] | 2023 | CS | Centre | Community | Probabilistic | Community health worker | Low | 32 | 15.62 | 68.75 | 68.75 | NR |
| Tchamani,2023 [47] | 2023 | CS | Centre | Community | Non-probabilistic | Women | Low | - | 75/156=48.08 | NR | NR | 4/239 = 4.18 |
| Njoh,2024 [48] | 2024 | CS | Multi-region | Online | Non-probabilistic | Healthcare worker | Low | 1225 | NR | 83.02 | NR | NR |
| Ntonifor,2024 [13] | 2024 | CS | South-West | Community | Non-probabilistic | Parent of eligible children | Low | 1187 | NR | 53.24 | NR | NR |
| Kehbila,2025 [49] | 2025 | CS | Centre | Community | Probabilistic | Parent of eligible children | Low | 397 | NR | 67.51 | NR | 29.97 |

1: Year of study completion; 2: Prevalence of individuals who are aware of the HPV vaccine that reduce the risk of uterine cervical cancers occurrence; 3: Prevalence of individuals who were willing to vaccinate or get their relative vaccinated if a HPV vaccine were freely made available for them ; 4: Prevalence of individuals who can recommend other people to get vaccinated; NR: Not reported; CS: Cross-sectional; RCT: Randomized control trial;

### 3.3. Willingness to vaccinate against human papillomavirus

The reported pooled prevalence of willingness to vaccinate targeted children against HPV in Cameroon was 68.08% (95% CI: 57.39-77.16; 12 reports; n = 4,993 participants), with high heterogeneity observed between studies (*I*^2^ = 98.0; *p* ˂ 0.001) (**Fig. 2**).

**Fig. 2.**
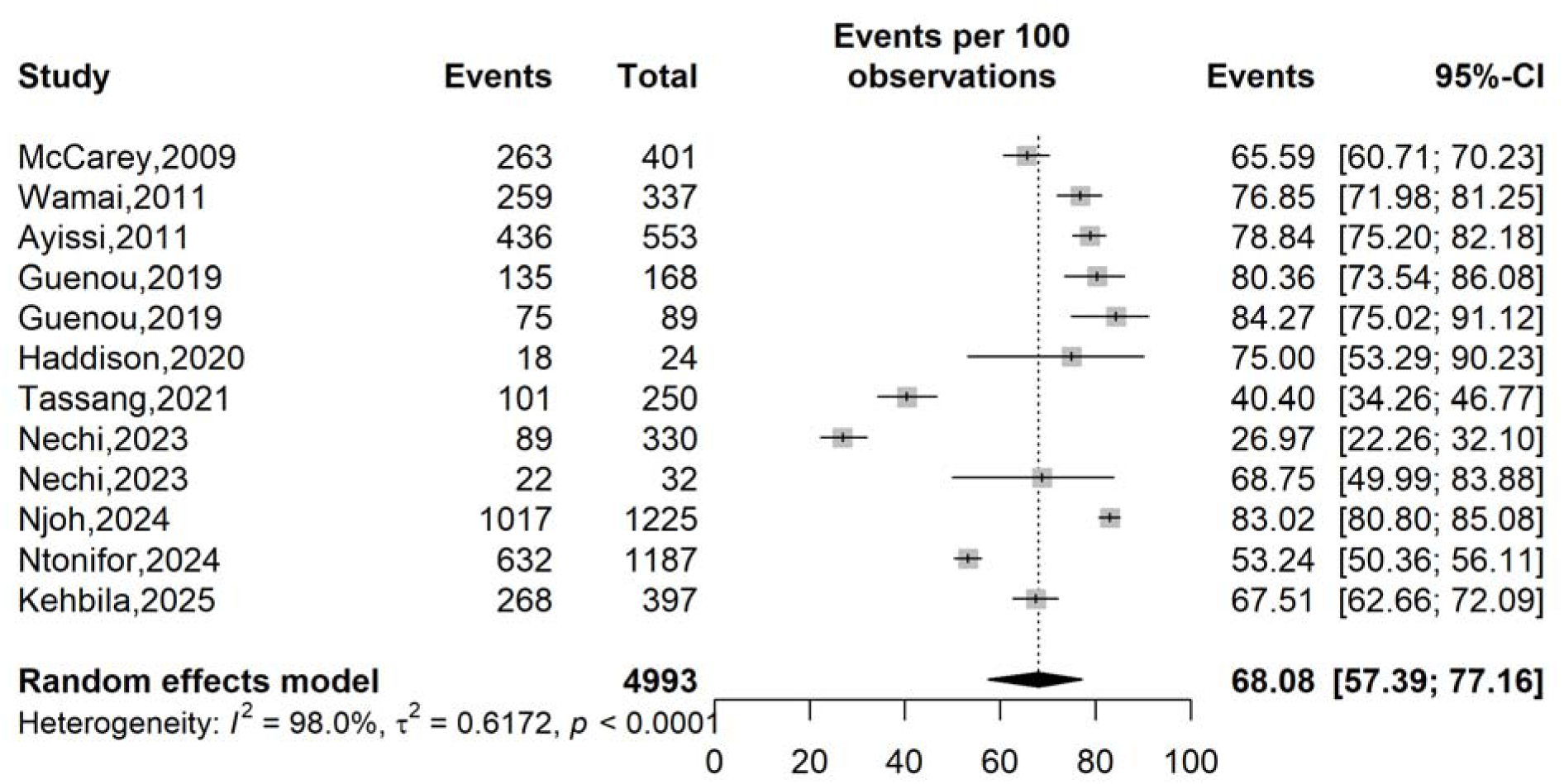
Forest plot displaying the pooled prevalence of willingness to vaccinate against the human papillomavirus in Cameroon.

Significant heterogeneity was observed across study timeframes (*p* = 0.006), settings (*p* ˂ 0.001), study regions (*p* ˂ 0.001), and participants type (*p* ˂ 0.001). The prevalence of willingness to vaccinate significantly declined (*p* = 0.006) over time, from 74.15% (95% CI: 67.19-80.07; 3 studies; n = 1,291) before 2014 to 57.50% (95% CI: 40.44-72.93; 6 studies; n = 3,421). Community-based studies reported significantly lower prevalence of willingness to vaccinate (55.72%; 95% CI: 40.55-69.90; 6 studies; n = 2,533) compared to other settings (Hospital: 76.49%; 95% CI: 67.47-83.62; 4 studies; n = 682). Regarding participant type, significantly higher prevalences of willingness to vaccinate were observed in studies conducted among healthcare workers (77.79%; 95% CI: 68.63-84.87; 4 studies; n = 1,739) and parents of eligible children (62.03%, 95%CI: 43.34-77.73; 5 studies; n = 2,419) (**Table 2 and Supplementary Fig. 1-5**).

**Table 2.** Subgroup meta-analysis of the pooled prevalence of willingness to vaccinate against the human papillomavirus in Cameroon.

| Subgroup | Sample size | Prevalence (%) | 95% CI limits<sup>1</sup> |  | Number of studies | Heterogeneity statistic<sup>1</sup> |  | Subgroup difference p-value |
| --- | --- | --- | --- | --- | --- | --- | --- | --- |
|  |  |  | Lower | Upper |  | I<sup>2</sup>(%) | p-value |  |
| **Study period (year)<sup>2</sup>** |  |  |  |  |  |  |  | 0.006 |
| □ 2014 | 1,291 | 74.15 | 67.19 | 80.07 | 3 | 91.2 | □ 0.001 |  |
| 2014-2020 | 281 | 81.14 | 76.14 | 85.30 | 3 | 0.0 | 0.544 |  |
| 2021-2023 | 3,421 | 57.50 | 40.44 | 72.93 | 6 | 98.9 | □ 0.001 |  |
| **Study setting** |  |  |  |  |  |  |  | □ 0.001 |
| Community | 2,533 | 55.72 | 40.55 | 69.90 | 6 | 97.5 | □ 0.001 |  |
| Hospital | 682 | 76.49 | 67.47 | 83.62 | 4 | 85.1 | □ 0.001 |  |
| Online | 1,225 | 83.02 | 80.80 | 85.08 | 1 | - | - |  |
| School | 553 | 78.84 | 75.20 | 82.18 | 1 | - | - |  |
| **Region** |  |  |  |  |  |  |  | □ 0.001 |
| Centre | 1,441 | 67.88 | 52.97 | 79.87 | 7 | 96.9 | □ 0.001 |  |
| North-West | 890 | 78.09 | 75.25 | 80.69 | 2 | 0.0 | 0.487 |  |
| South-West | 1,438 | 47.39 | 38.59 | 56.36 | 2 | 92.6 | □ 0.001 |  |
| Multiregion | 1,225 | 83.02 | 80.80 | 85.08 | 1 | - | - |  |
| **Sampling method** |  |  |  |  |  |  |  | 0.139 |
| Non-probabilistic | 4,234 | 72.16 | 62.31 | 80.25 | 9 | 97.9 | □ 0.001 |  |
| Probabilistic | 759 | 53.68 | 30.57 | 75.31 | 3 | 98.3 | □ 0.001 |  |
| **Participant** |  |  |  |  |  |  |  | □ 0.001 |
| Women from the general population | 250 | 40.40 | 34.26 | 46.77 | 1 | - | - |  |
| Healthcare worker | 1,739 | 77.79 | 68.63 | 84.87 | 4 | 94.5 | □ 0.001 |  |
| Parent<sup>4</sup> | 2,419 | 62.03 | 43.34 | 77.73 | 5 | 98.1 | □ 0.001 |  |
| Community health worker | 32 | 68.75 | 49.99 | 83.88 | 1 | - | - |  |
| Adolescent girl | 553 | 78.84 | 75.20 | 82.18 | 1 | - | - |  |

<sup>1</sup> Random effects model; <sup>2</sup> □ 2014 = Pre-HPV vaccination in Cameroon, 2014-2020= HPV vaccination demonstration in Cameroon, 2021-2023= post-HPV vaccine introduction in Cameroon; <sup>3</sup> Parent of HPV vaccine eligible children

### 3.4. Awareness of the preventive vaccine against cervical cancer

The pooled prevalence of HPV vaccine awareness as preventive method against cervical cancer in Cameroon was 41.29% (95% CI: 28.74-55.08; 33 studies; n = 8,175 participants) with high heterogeneity across included studies (*I*^2^ = 97.9; *p* ˂ 0.001) (**Fig. 3**).

**Fig. 3.**
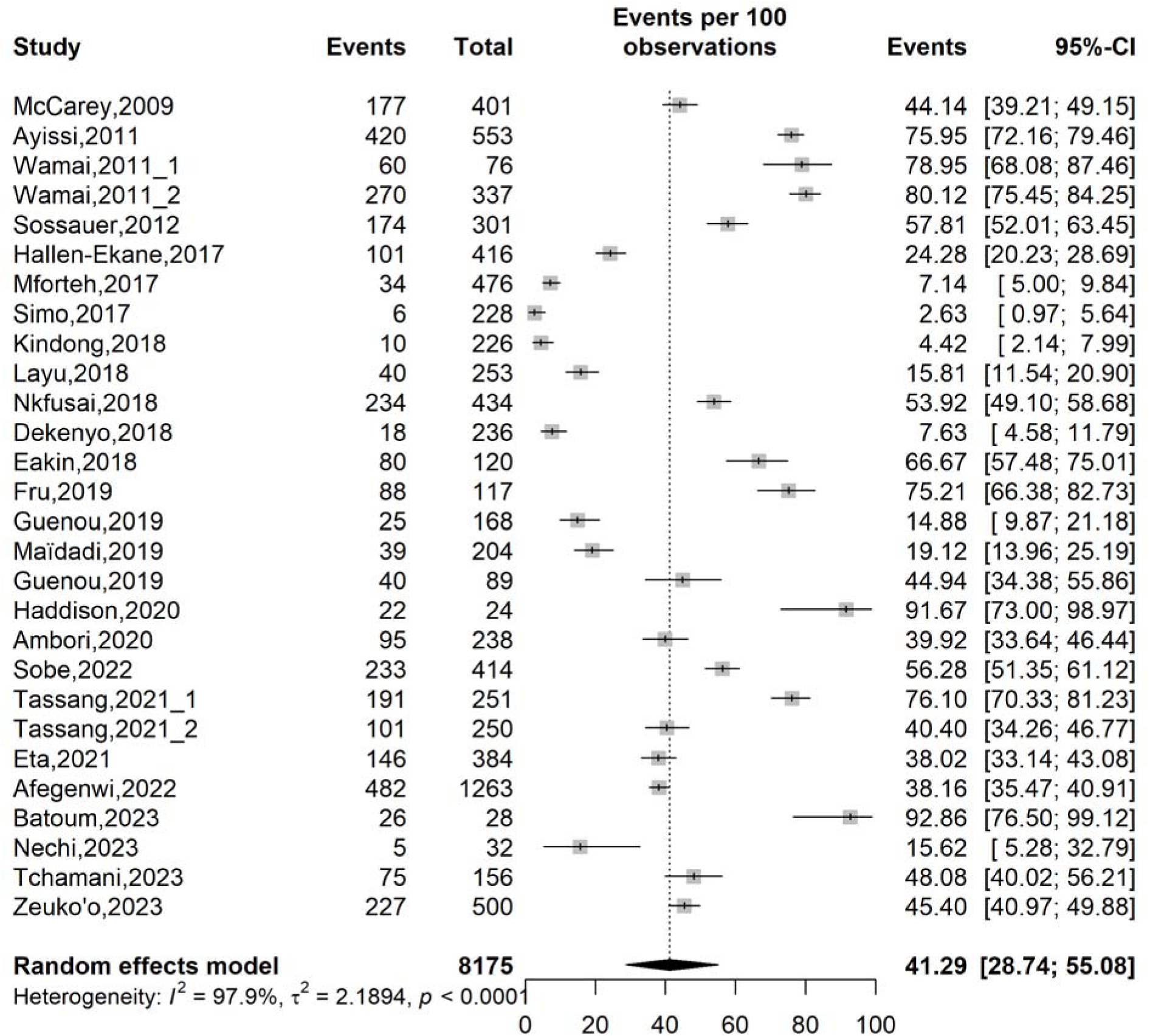
Forest plot displaying the pooled prevalence of human papillomavirus-vaccine awareness in Cameroon.

Study timeframe (*p* = 0.002), study regions (*p* ˂ 0.001), sampling method (*p* ˂ 0.001), and participants type (*p* ˂ 0.001) were significant sources of heterogeneity in HPV vaccine awareness estimates. Awareness declined over time with a significant drop observed during the 2014-2020 period (95% CI: 13.33-45.62; 14 studies; n = 3,229) compared to higher estimates were recorded before 2014 (68.49%; 95% CI: 55.12-79.38; 5 studies; n = 1,668). The study revealed significant regional disparities across regions; the lowest awareness observed in the West region (4.73%; 95% CI: 2.23-9.75; 2 studies; n = 464) while higher rates were found in the Centre region (48.02%; 95% CI: 26.44-70.37; 9 studies; n = 1,403); South-West (49.98%; 95% CI: 38.94-61.01; 8 studies; n = 2,489) and Far North (76.10%; 95% CI: 70.33-81.23; 1 study; n = 251). Regarding the type of participants, the highest awareness rate significantly was observed among healthcare workers (74.49%; 95% CI: 49.46-89.71; 5 studies; n = 618) and students (51.11%; 95% CI: 36.34-65.68; 6 studies; n = 3,266), while the lowest rates were identified among women from the general population (27.05%; 95% CI: 13.93-45.93; 13 studies; n = 3,370) and community health workers (15.62%; 95% CI: 5.28-32.79; 1 study; n = 32) (**Table 3 and Supplementary Fig. 6-10**).

**Table 3.**
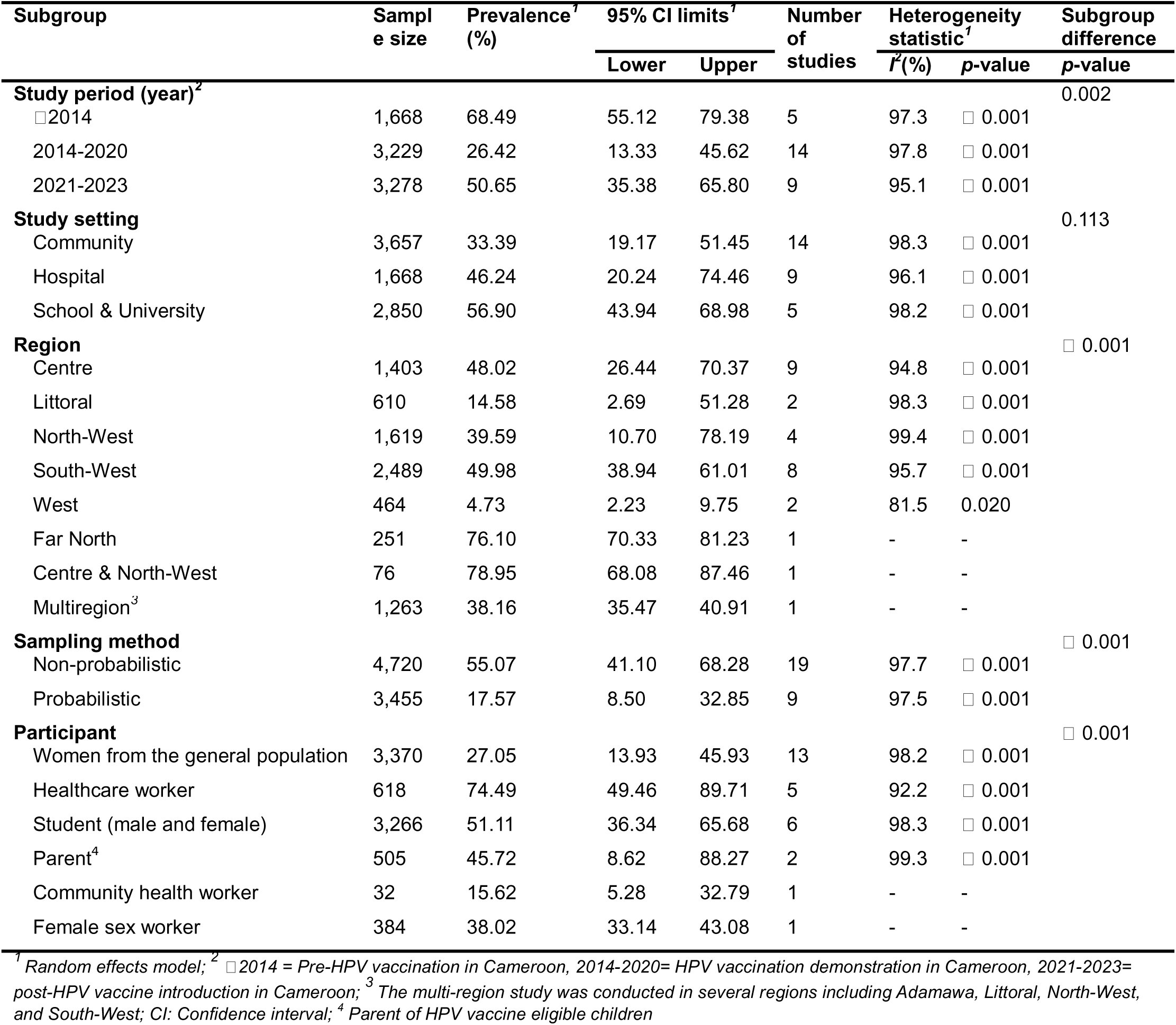
Subgroup meta-analysis of the pooled prevalence of human papillomavirus-vaccine awareness in Cameroon.

| Subgroup | Sample size | Prevalence<sup>1</sup> (%) | 95% CI limits<sup>1</sup> |  | Number of studies | Heterogeneity statistic<sup>1</sup> |  | Subgroup difference p-value |
| --- | --- | --- | --- | --- | --- | --- | --- | --- |
|  |  |  | Lower | Upper |  | I<sup>2</sup>(%) | p-value |  |
| **Study period (year)<sup>2</sup>** |  |  |  |  |  |  |  | 0.002 |
| □ 2014 | 1,668 | 68.49 | 55.12 | 79.38 | 5 | 97.3 | □ 0.001 |  |
| 2014-2020 | 3,229 | 26.42 | 13.33 | 45.62 | 14 | 97.8 | □ 0.001 |  |
| 2021-2023 | 3,278 | 50.65 | 35.38 | 65.80 | 9 | 95.1 | □ 0.001 |  |
| **Study setting** |  |  |  |  |  |  |  | 0.113 |
| Community | 3,657 | 33.39 | 19.17 | 51.45 | 14 | 98.3 | □ 0.001 |  |
| Hospital | 1,668 | 46.24 | 20.24 | 74.46 | 9 | 96.1 | □ 0.001 |  |
| School & University | 2,850 | 56.90 | 43.94 | 68.98 | 5 | 98.2 | □ 0.001 |  |
| **Region** |  |  |  |  |  |  |  | □ 0.001 |
| Centre | 1,403 | 48.02 | 26.44 | 70.37 | 9 | 94.8 | □ 0.001 |  |
| Littoral | 610 | 14.58 | 2.69 | 51.28 | 2 | 98.3 | □ 0.001 |  |
| North-West | 1,619 | 39.59 | 10.70 | 78.19 | 4 | 99.4 | □ 0.001 |  |
| South-West | 2,489 | 49.98 | 38.94 | 61.01 | 8 | 95.7 | □ 0.001 |  |
| West | 464 | 4.73 | 2.23 | 9.75 | 2 | 81.5 | 0.020 |  |
| Far North | 251 | 76.10 | 70.33 | 81.23 | 1 | - | - |  |
| Centre & North-West | 76 | 78.95 | 68.08 | 87.46 | 1 | - | - |  |
| Multiregion<sup>3</sup> | 1,263 | 38.16 | 35.47 | 40.91 | 1 | - | - |  |
| **Sampling method** |  |  |  |  |  |  |  | □ 0.001 |
| Non-probabilistic | 4,720 | 55.07 | 41.10 | 68.28 | 19 | 97.7 | □ 0.001 |  |
| Probabilistic | 3,455 | 17.57 | 8.50 | 32.85 | 9 | 97.5 | □ 0.001 |  |
| **Participant** |  |  |  |  |  |  |  | □ 0.001 |
| Women from the general population | 3,370 | 27.05 | 13.93 | 45.93 | 13 | 98.2 | □ 0.001 |  |
| Healthcare worker | 618 | 74.49 | 49.46 | 89.71 | 5 | 92.2 | □ 0.001 |  |
| Student (male and female) | 3,266 | 51.11 | 36.34 | 65.68 | 6 | 98.3 | □ 0.001 |  |
| Parent<sup>4</sup> | 505 | 45.72 | 8.62 | 88.27 | 2 | 99.3 | □ 0.001 |  |
| Community health worker | 32 | 15.62 | 5.28 | 32.79 | 1 | - | - |  |
| Female sex worker | 384 | 38.02 | 33.14 | 43.08 | 1 | - | - |  |

<sup>1</sup> Random effects model; <sup>2</sup> □ 2014 = Pre-HPV vaccination in Cameroon, 2014-2020= HPV vaccination demonstration in Cameroon, 2021-2023= post-HPV vaccine introduction in Cameroon; <sup>3</sup> The multi-region study was conducted in several regions including Adamawa, Littoral, North-West, and South-West; CI: Confidence interval; <sup>4</sup> Parent of HPV vaccine eligible children

### 3.5. Human papillomavirus vaccine recommendation and uptake

This study showed that in Cameroon, 67,67% (95% CI: 50.67-81.01; 8 studies; n = 1,617) of individuals recommended vaccination against HPV to other community members; with high heterogeneity across studies (*I*^2^ = 97.3%; *p* ˂ 0.001). However, A much lower proportion of the population reported having ever actually received the HPV vaccine (22.92%; 95% CI: 6.89-54.46; 9 studies; n = 9,686) with similar with high heterogeneity between studies (*I*^2^ = 99.7%; *p* ˂ 0.001) (**Fig 4 and 5**).

**Fig. 4.**
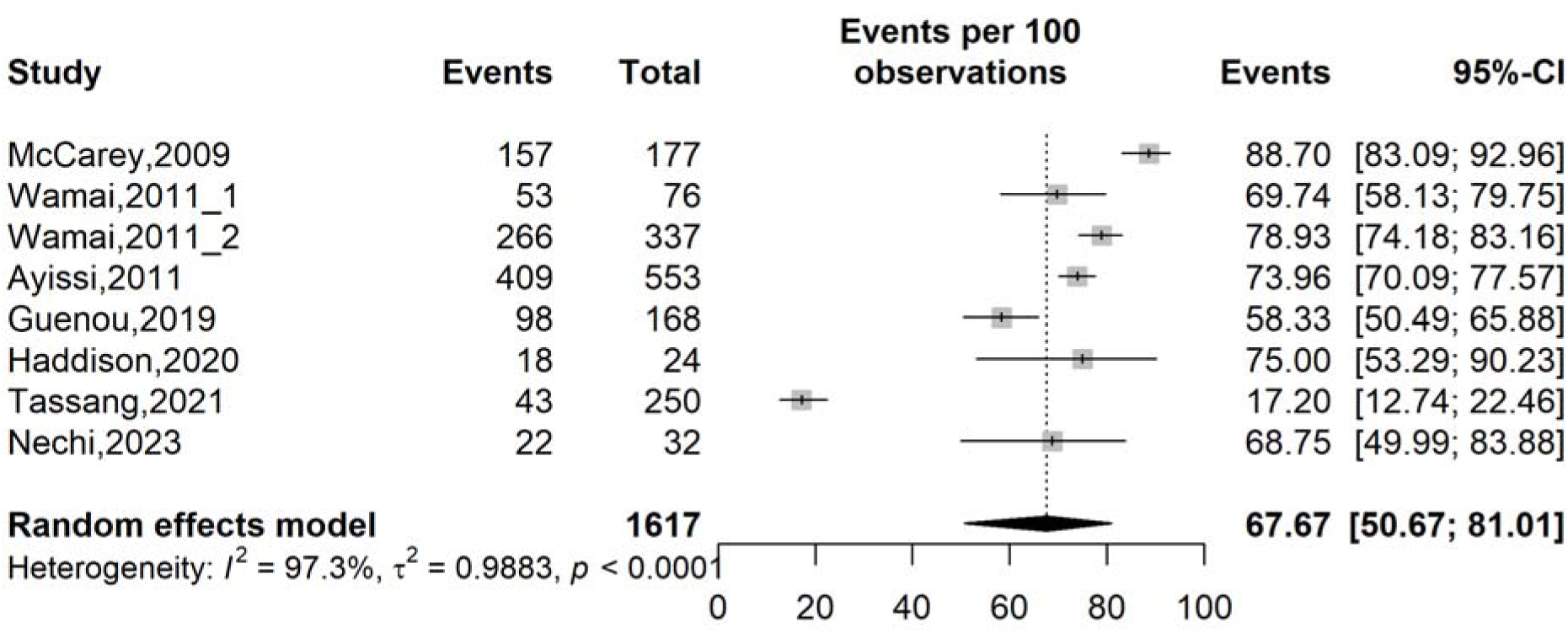
Forest plot displaying the pooled prevalence of human papillomavirus-vaccine recommendation in Cameroon.

**Fig. 5.**
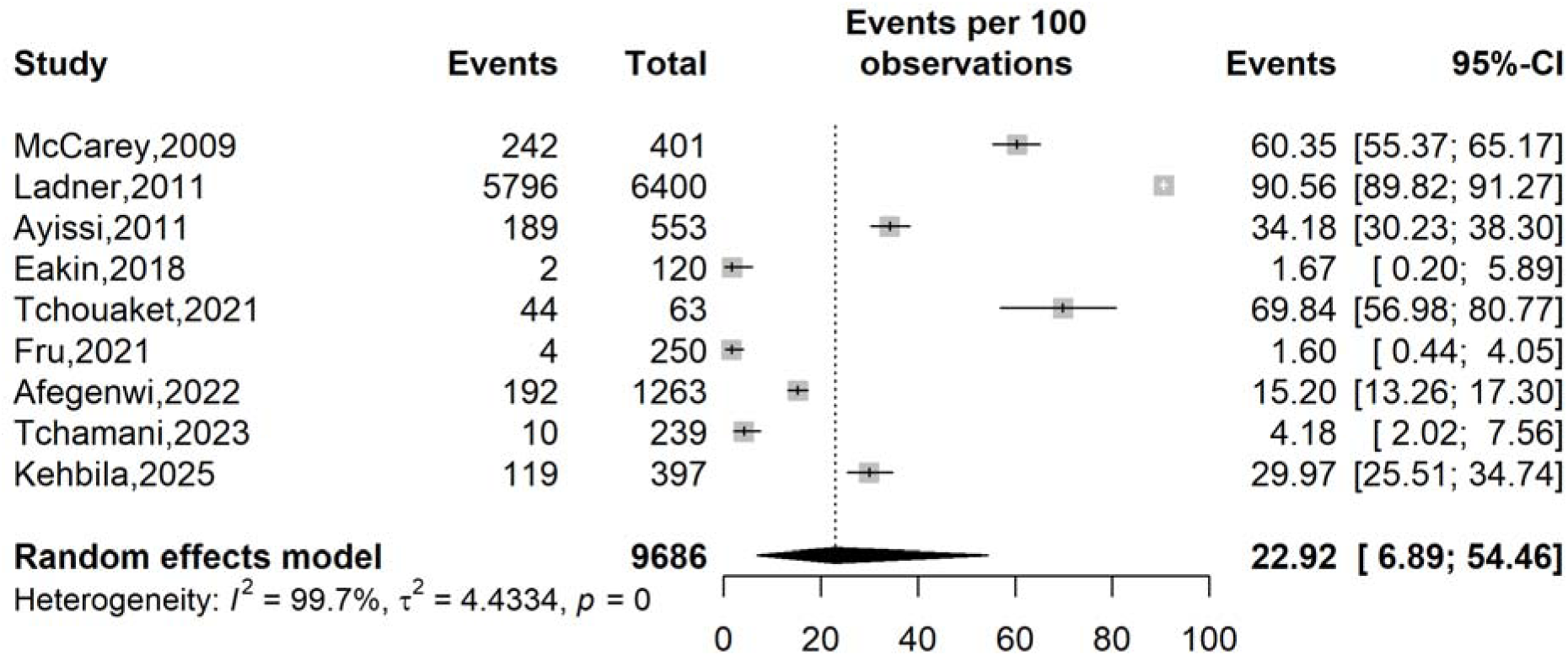
Forest plot displaying the pooled prevalence of human papillomavirus-vaccine vaccine uptake in Cameroon.

### 3.6. Publication bias and sensitivity analysis

There was a slight asymmetry in the funnel plot assessing the risk of publication among studies assessing the prevalence of willingness to vaccinate against HPV. However, the Egger’s (*p* = 0.707) and Begg’s (*p* = 0.784) tests revealed non-significant risk of publication bias for this estimate (**Supplementary Fig. 11**). The sensitivity analysis showed no study significantly influenced the pooled prevalence of willingness to vaccinate against HPV (**Supplementary Fig. 12**).

For the pooled prevalence of HPV vaccine awareness estimates, a symmetrical funnel plot was observed suggesting low risk of publication bias; the Egger’s (*p* = 0.455) and Begg’s (*p* = 0.323) tests further confirmed this conclusion (**Supplementary Fig. 13**). Moreover, no single study significantly influenced the resulting pooled estimate (**Supplementary Fig. 14**).

Sensitivity analysis of the pooled HPV vaccine recommendation showed that no single study significantly influenced the overall estimate, confirming the robustness of the findings. Although, the funnel plot appeared slightly asymmetrical, the trim and fill analysis identified no missing studies, suggesting an absence of publication bias (**Supplementary Fig. 15-17**).

In contrast, sensitivity analysis for the HPV vaccine uptake showed the significant influence of some included studies except the study conducted by Ladner *et al.* [23]. By omitting these studies one at a time, the pooled estimates increased from 23% to 67-71%. The funnel plot assessing the risk of publication bias showed asymmetry suggesting risk of publication bias and the trim and fill analysis imputed five potential unpublished studies and yielded a prevalence of HPV vaccine uptake of 78.71% (95% CI: 33.03-96.52; *I*^2^ = 99.8%; *p* ˂ 0.001). This suggested that the exact overall pooled estimate of HPV vaccine uptake might be higher than that obtained in this study suggesting that our findings should be interpreted with caution (**Supplementary Fig. 18-20**).

### 3.7. Determinants of human papillomavirus vaccine hesitancy

Individuals with no knowledge of HPV were nearly three times more likely to be hesitant toward HPV vaccination compared to their counterparts (OR: 2.58; 95% CI: 2.06-3.22); 2 studies; *I*^2^ = 0.0; *p* = 0.606). Although not statistically significant, female gender (OR: 1.18; 95% CI: 0.82-1.69; 3 studies; *I*^2^ = 63.7; *p* = 0.064) and lack of knowledge regarding cervical cancer (OR: 7.97; 95% CI: 0.46-137.83; 2 studies; *I*^2^ = 97.6; *p* ˂ 0.001) Were associated with an 18% and nearly eight-fold increase in the likelihood of hesitancy, respectively (**Table 4 and Supplementary Fig. 21-23**).

**Table 4.** Determinants of human papillomavirus vaccine hesitancy in Cameroon.

| Predictor | Modality | OR <sup>1</sup> | 95% CI |  | Reports included | Heterogeneity |  |
| --- | --- | --- | --- | --- | --- | --- | --- |
|  |  |  | Lower | Upper |  | I<sup>2</sup> (%) | p-value |
| **Gender** | Female vs. Male | 1.18 | 0.82 | 1.6 | 3 | 63.7 | 0.064 |
| **Knowledge of HPV** | No vs. Yes | 2.58 | 2.06 | 3.22 | 2 | 0.0 | 0.606 |
| **Knowledge of cervical cancer** | No vs. Yes | 7.97 | 0.46 | 137.83 | 2 | 97.6 | □ 0.001 |

<sup>1</sup>Random effect model; OR: Odds ratio; CI: Confidence interval; HPV: Human papillomavirus

## 4. Discussions

This meta-analysis offers a comprehensive synthesis of the current evidence on HPV vaccine awareness, willingness to vaccinate, and actual uptake of this vaccine in Cameroon, revealing a disconnect between intention and practice. While parental willingness to vaccinate children above average, overall awareness of the HPV vaccine remains insufficient, and actual vaccination coverage was low.

### 4.1. Willingness to vaccinate against human papillomavirus

The pooled prevalence of parental willingness to vaccinate targeted children against HPV in Cameroon was 68% (95% CI: 57–77), with high heterogeneity. Similar findings were reported in a meta-analysis conducted in Ethiopia, where the acceptance rate reached 72% (95% CI: 58–86%) [50]. This consistency may reflect the strong prioritization of children’s health within many communities, where vaccination is perceived as a means of protecting daughters’ future health and fertility. This favorable perception is likely reinforced by the persistently high burden of HPV infection in the region [10, 50, 51]. Such parental and healthcare professionals’ compliance with this vaccine represents a critical opportunity for public health policymakers to strengthen the implementation and scale-up of HPV vaccination, particularly through school and community-based delivery strategies.

The prevalence of willingness to vaccinate significantly declined over time, decreasing from 74% before 2014 to 57% thereafter. This decline may reflect the growing influence of misinformation, the spread of rumors related to vaccine safety, and fluctuating levels of trust in vaccination programs [13]. Furthermore, the HPV vaccination campaign, launched in October 2020 during the COVID-19 pandemic, occurred within a context of widespread vaccine hesitancy. This environment likely exacerbated fears and facilitated the dissemination of misinformation surrounding the HPV vaccine [36].

Studies conducted in community settings reported a significantly lower prevalence of vaccine acceptability (56%) compared with those carried out in hospital settings (76%). This disparity may be explained by more limited access to reliable health information in community environments, which may promote misinformation and the persistence of sociocultural beliefs unfavorable to vaccination [52]. Conversely, hospital-based participants benefit from closer interaction with healthcare professionals, greater exposure to awareness campaigns, and trusted source of information from healthcare providers, thereby enhancing vaccine acceptability [53].

Regarding the type of participants, the willingness to vaccinate was particularly high among healthcare professionals (78%), reflecting their level of scientific knowledge, their perception of the burden of HPV-related diseases, as well as their confidence in the efficacy and safety of the vaccines [44].

### 4.2. Awareness of the preventive vaccine against cervical cancer

The present meta-analysis estimated a pooled HPV vaccine awareness prevalence of 41% (95% CI: 29–55) in Cameroon, with high heterogeneity. This insufficient level of awareness is likely multifactorial. Health system constraints, such as staff shortages, heavy workloads, and geographic barriers, impede access to information and vaccination services. The dissemination of information is also hindered by sociocultural factors, including perceptions linking HPV vaccination to sexuality, taboos surrounding reproductive health discussions, and reluctance from some community leaders [9, 54, 55].

Our analysis also revealed marked regional disparities in HPV vaccine knowledge in Cameroon, with extremely low awareness in the West region (4.73%) compared with the Centre (48.02%), South-West (49.98%), and Far North (76.10%) regions. In the Centre region, a high density of healthcare infrastructure and close integration with national vaccination initiatives have improved access to trustworthy information and the dissemination of preventive messages [56]. These efforts are reinforced by regular awareness campaigns, pilot projects in health facilities, and intensified media engagement designed to counter HPV vaccine-related misinformation [57, 58]. In the South-West, community-based approaches involving schools, religious leaders, and community health workers appear to enhance knowledge despite structural healthcare constraints [31]. Conversely, the very low awareness observed in the West region may result from limited targeted awareness initiatives and persistent sociocultural beliefs unfavorable to vaccination, combined with reliance on informal information sources [59]. The unexpectedly high knowledge level reported in the Far North region may reflect targeted interventions in underserved areas, including community mobilization and proximity-based awareness campaigns, although this finding should be interpreted cautiously due to the limited number of studies [9]. The observed high heterogeneity highlights the need for context-specific regional strategies rather than uniform national approaches to sustainably improve HPV vaccine awareness and acceptability in Cameroon.

Regarding participant type, marked disparities were observed, with awareness highest among healthcare workers (74%) and students (51%), and considerably lower among women from the general population (27%) and community health workers (15%). Elevated awareness among healthcare workers likely reflects professional training and continuous exposure to scientific information and immunization programs [24, 44]. Students may likewise benefit from school-based vaccination initiatives and greater access to digital information channels [10, 45]. Conversely, the limited awareness among community health workers points to insufficient training and a predominant focus on operational responsibilities rather than comprehensive health education, despite their critical intermediary role between health services and communities [36, 48]. Lower awareness among women from the general population further highlights structural inequities in access to preventive information, particularly in rural and peri-urban settings [60]. Similar trends reported across Ethiopia, Nigeria, and Kenya underscore persistent population-level informational gaps across sub-Saharan Africa [61–63]. Collectively, these findings emphasize the need to strengthen community health worker capacity and implement context-specific communication strategies alongside sustained school-based interventions.

A significant variation in awareness rates was also observed depending on the study period. A notable decline in awareness was observed over time, with a significant drop during the 2014-2020 period (26%), compared to the higher estimates recorded before 2014 (68%). The decline in HPV awareness observed after 2014 likely reflects both programmatic and methodological factors. Early estimates coincided with pilot HPV vaccination initiatives characterized by intensive community sensitization and educational interventions, which were shown to substantially improve knowledge levels [8, 23]. Following program integration into routine health services, communication activities appear to have decreased, with greater emphasis placed on operational delivery rather than sustained education [36, 48]. Furthermore, later studies increasingly included community-based and rural populations with limited exposure to health information.

### 4.3. Vaccine recommendation and uptake

In this meta-analysis, more than two-thirds of the population (67%) reported recommending HPV vaccination to their relatives and peers, only 23% indicated having actually received the vaccine, highlighting a discrepancy between supportive attitudes and effective uptake. This gap illustrates a well-recognized intention behavior divide in low-resource settings, primarily driven by persistent structural constraints, including limited vaccine availability, geographical barriers to healthcare access, and indirect costs such as transportation expenses and time required to obtain services. Beyond access-related barriers, individual and perceptual determinants further contribute to low vaccine uptake [64, 65]. Many young women, who mistakenly perceive themselves as being at low risk of cervical cancer because of their young age, may deprioritize vaccination despite expressing favorable views toward immunization [47]. In addition, ongoing security crises in the North-West, South-West, and Far North regions have substantially disrupted healthcare delivery, further restricting access to vaccination services and exacerbating structural inequities related to geographic isolation and insecurity [9].

Individuals with no prior knowledge of HPV were nearly three times more likely to exhibit hesitancy toward HPV vaccination compared with their informed counterparts. This finding may be explained by the fact that individuals lacking awareness of HPV often underestimate the risks associated with the virus and are more susceptible to misinformation and rumors, which can generate doubts about vaccine effectiveness and safety, ultimately increasing reluctance toward vaccination compared with better-informed individuals [47].

## 5. Limitations

This review has several limitations. Substantial heterogeneity was observed across included studies, which may limit the generalizability of pooled estimates despite subgroup analyses. Many studies relied on non-probabilistic sampling methods, increasing the risk of selection bias. In addition, the use of self-reported data may have introduced recall and social desirability biases, potentially leading to overestimation of outcomes. Geographic representation was uneven, with some regions underrepresented, and variations in survey instruments and definitions may have affected comparability.

## 6. Conclusions

This systematic review and meta-analysis synthesized existing evidence and contextualized previous findings within the evolving HPV vaccination landscape in Cameroon. Despite relatively favorable levels of awareness and willingness reported in some populations, actual vaccine uptake remains low, highlighting persistent gaps between intention and practice. The results reveal important regional and population disparities, reflecting inequities in access to information, preventive services, and vaccination opportunities. These findings emphasize the urgent need for coordinated, context-adapted strategies integrating health education, strengthened vaccination delivery systems, and community engagement to improve coverage. Policymakers and public health stakeholders should prioritize sustainable HPV prevention programs to reduce the burden of cervical cancer in Cameroon.

## Data Availability

All data generated or analyzed during this study are included in this published article and its supplementary data files.

## Declarations

### Ethics approval and consent to participate

Not applicable.

### Consent for publication

Non applicable.

### Author contributions

Conceptualization: FZLC; Data curation: FZLC, RT, and CA; Formal analysis: FZLC; Funding acquisition: None; Investigation: FZLC, RT, ATT, RT, EME and CA; Methodology: FZLC, RT, ATT, RT, EME and CA; Project administration: FZLC; Resources: All authors; Software: FZLC, RT, ATT, RT, EME and CA; Supervision: FZLC and INM; Validation: FZLC; Visualization: All authors; Writing – original draft: FZLC, RT, and CA; Writing – review and editing: All authors.

## Acknowledgements

None.

