## Supplementary Material for "Human papillomavirus vaccine acceptability in Cameroon: a systematic review and meta-analysis"

**Supplementary Table 1** Searching strategy by database

| **Database** | **Search string** | **Number of entries** |
| --- | --- | --- |
| **PubMed** | ("human papillomavirus"[TIAB] OR HPV[TIAB] OR "cervical cancer"[TIAB])  AND  (knowledge [TIAB])  AND  (Cameroon [Mesh] OR Cameroon [TIAB] OR Cameroonian [TIAB] OR Cameroun [TIAB]) | 57 |
| **Scopus** | TITLE-ABS-KEY ("human papillomavirus" OR HPV OR "cervical cancer")  AND  TITLE-ABS-KEY (vaccine OR vaccination OR vaccines OR knowledge OR awareness OR attitude OR practice)  AND  TITLE-ABS-KEY (Cameroon OR Cameroonian OR Cameroun) | 70 |
| **Web of sciences** | ("human papillomavirus" OR HPV OR "cervical cancer")  AND (vaccine OR vaccination OR vaccines OR knowledge OR awareness OR attitude OR practice)  AND  (Cameroon OR Cameroonian OR Cameroun) | 145 |
| **Embase** | ("human papillomavirus" OR HPV OR "cervical cancer")  AND (vaccine OR vaccination OR vaccines OR knowledge OR awareness OR attitude OR practice)  AND  (Cameroon OR Cameroonian OR Cameroun) | 83 |
| **Cochrane Library** | (“human papillomavirus” OR HPV OR “cervical cancer”)  AND  (vaccine OR vaccination OR knowledge OR awareness OR attitude OR practice)  AND  (Cameroon OR Cameroonian OR Cameroun) | 8 |
| **African Journals Online (AJOL)** | (“human papillomavirus” OR HPV OR “cervical cancer”)  AND  (vaccine OR vaccination OR knowledge OR awareness OR attitude OR practice)  AND  (Cameroon OR Cameroonian OR Cameroun) | 77 |
| **Health Sciences and Disease** | (“human papillomavirus” OR HPV OR “cervical cancer”)  AND  (vaccine OR vaccination OR knowledge OR awareness OR attitude OR practice)  AND  (Cameroon OR Cameroonian OR Cameroun) | 2 |

**Subgroup analysis**

**Study period**

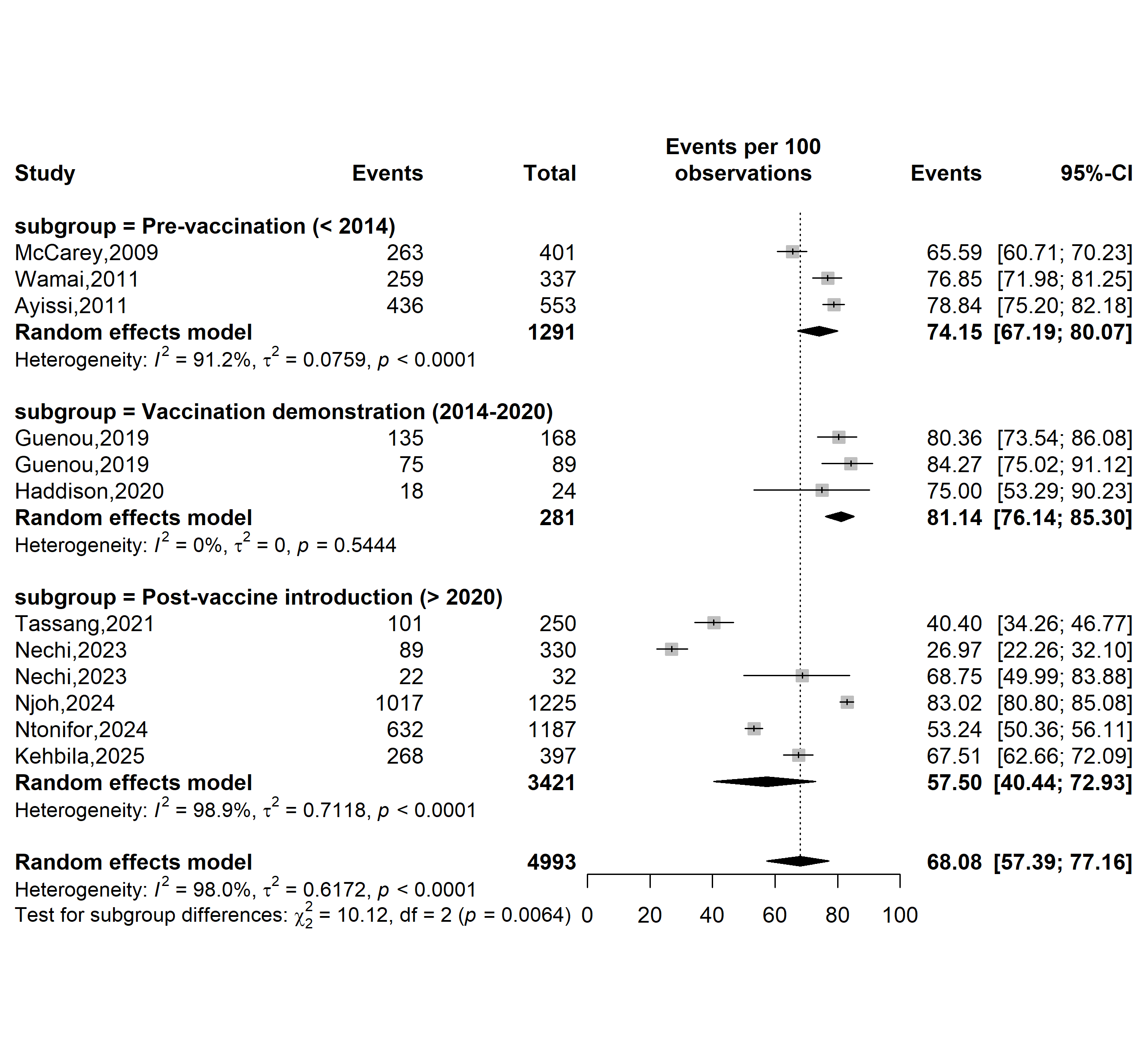

**Supplementary Fig. 1** Pooled prevalence of willingness to vaccinate against human papillomavirus (HPV) according to specific HPV vaccine introduction timeframe in Cameroon

**Study setting**

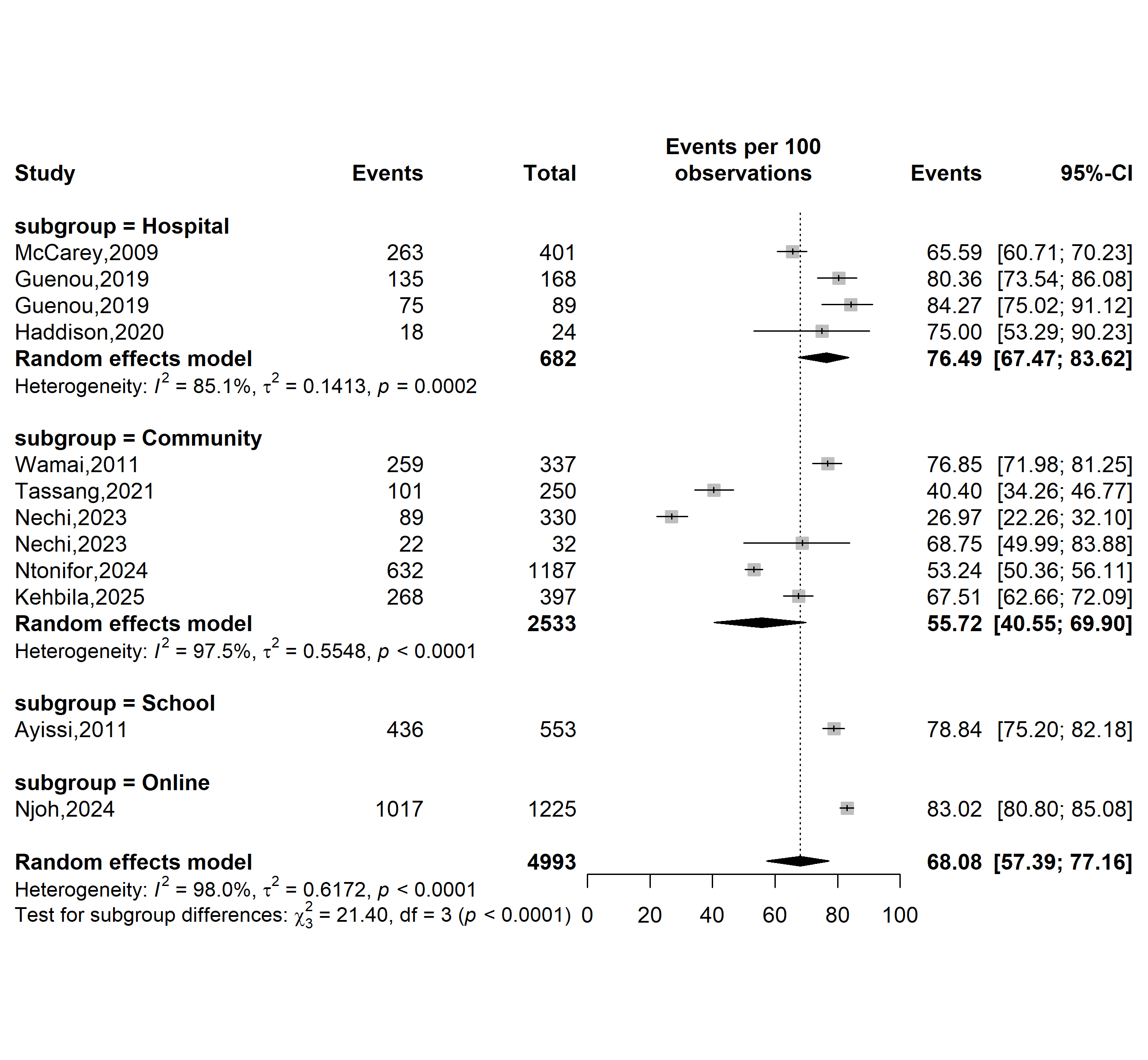

**Supplementary Fig. 2** Pooled prevalence of willingness to vaccinate against human papillomavirus according to study settings in Cameroon

**Study site**

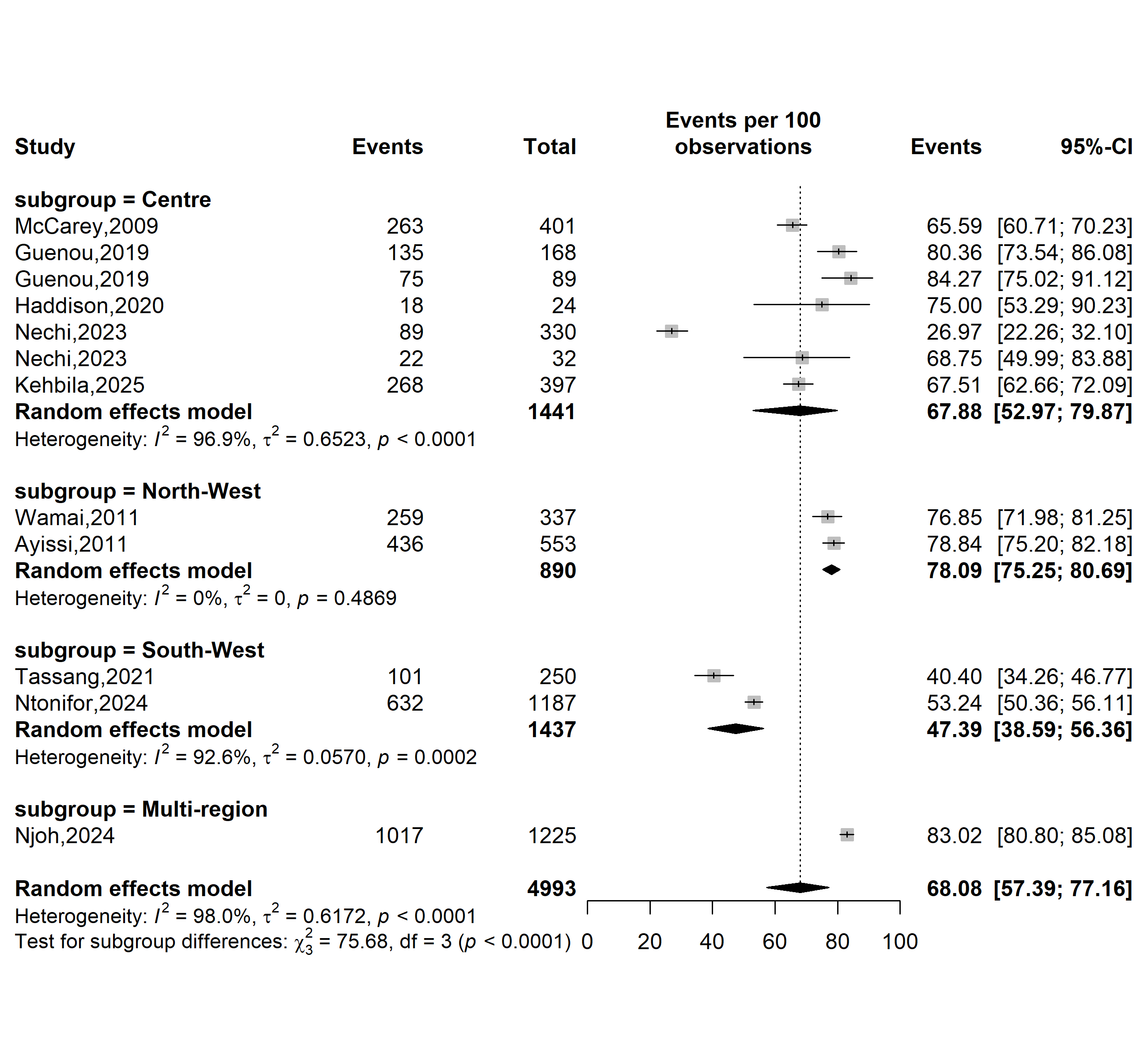

**Supplementary Fig. 3** Pooled prevalence of willingness to vaccinate against human papillomavirus according to study sites in Cameroon

**Sampling method**

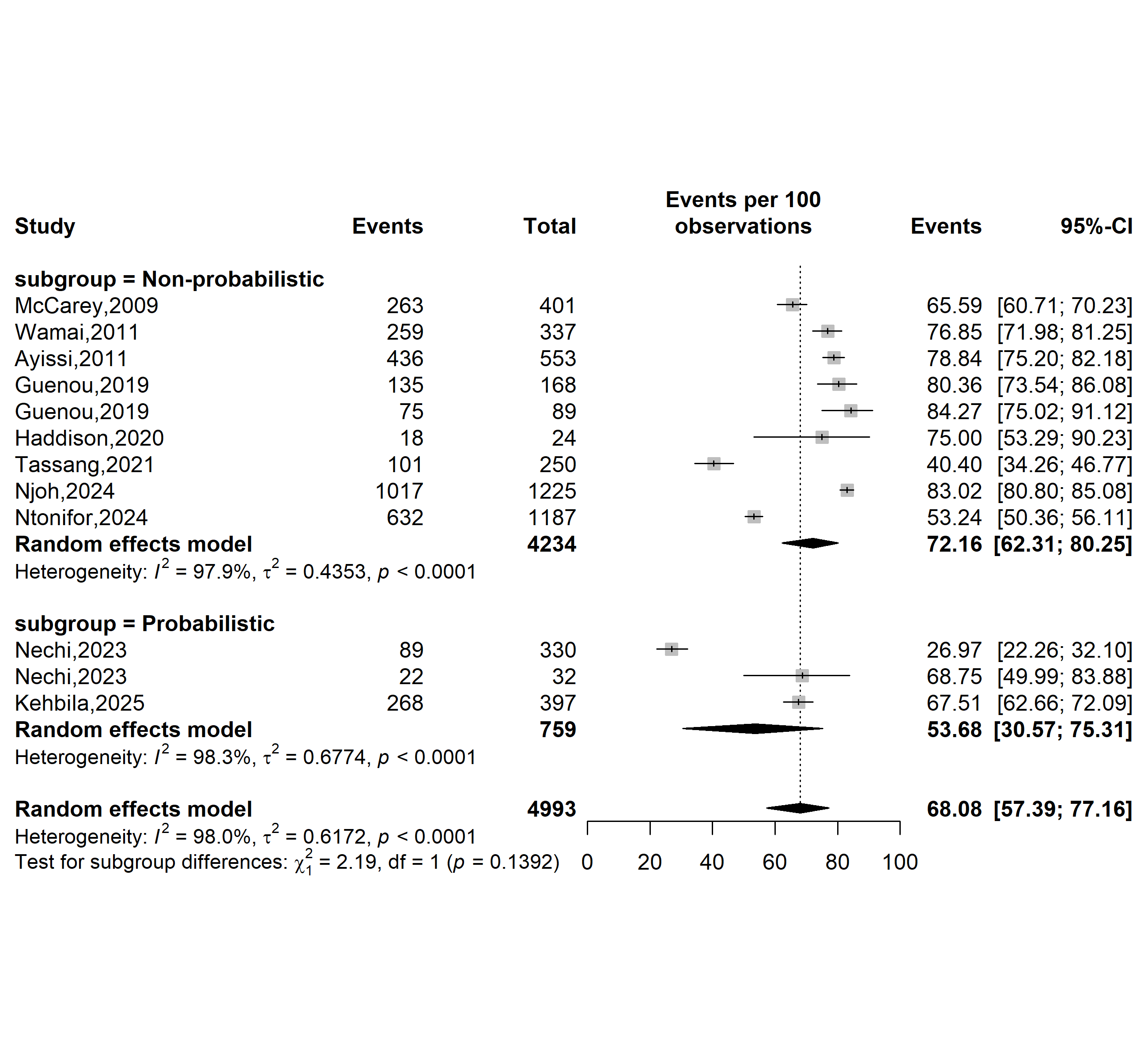

**Supplementary Fig. 4** Pooled prevalence of willingness to vaccinate against human papillomavirus by sampling methods in Cameroon

**Type of participants**

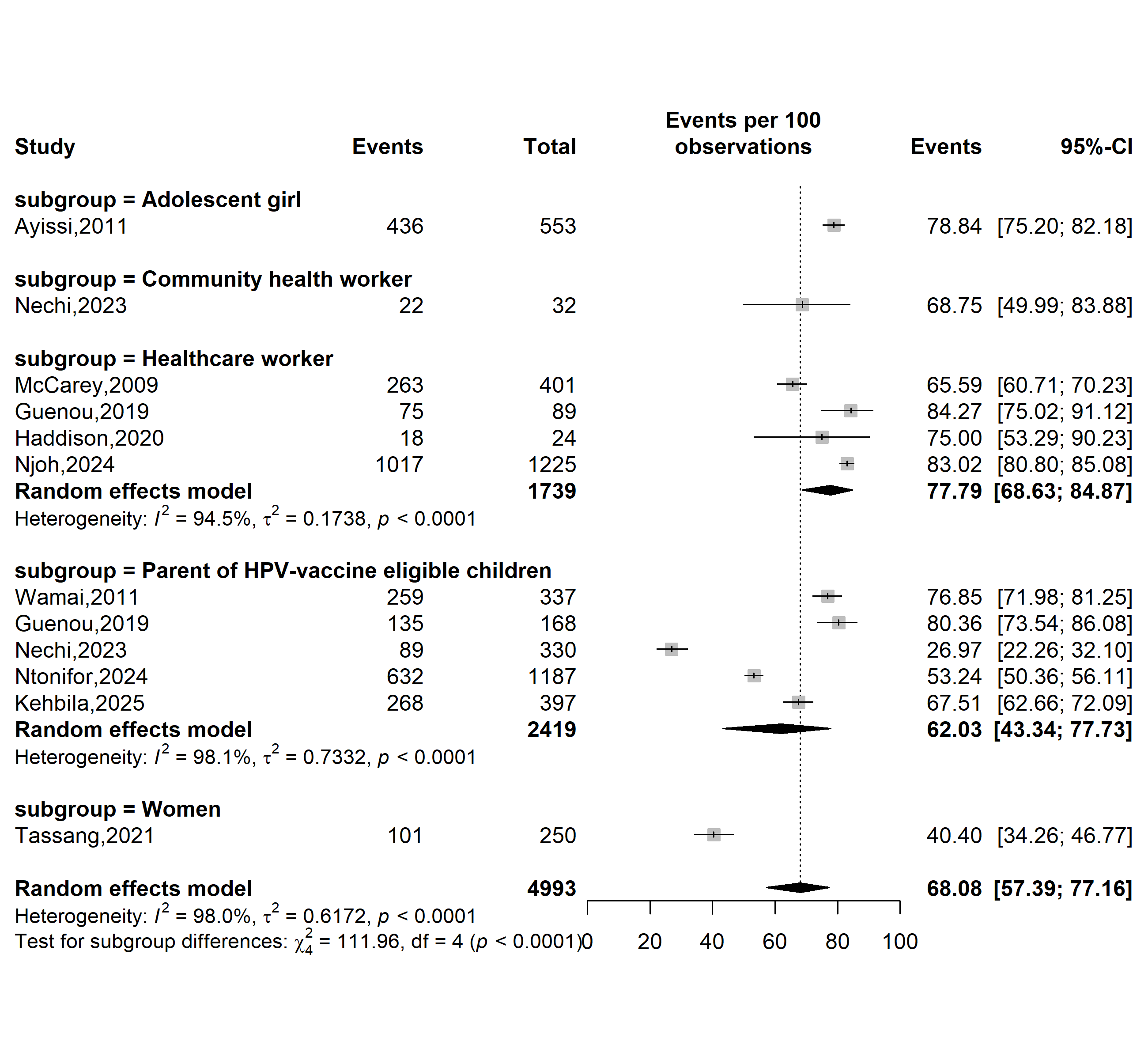

**Supplementary Fig. 5** Pooled prevalence of willingness to vaccinate against human papillomavirus by sampling methods in Cameroon

**Study period**

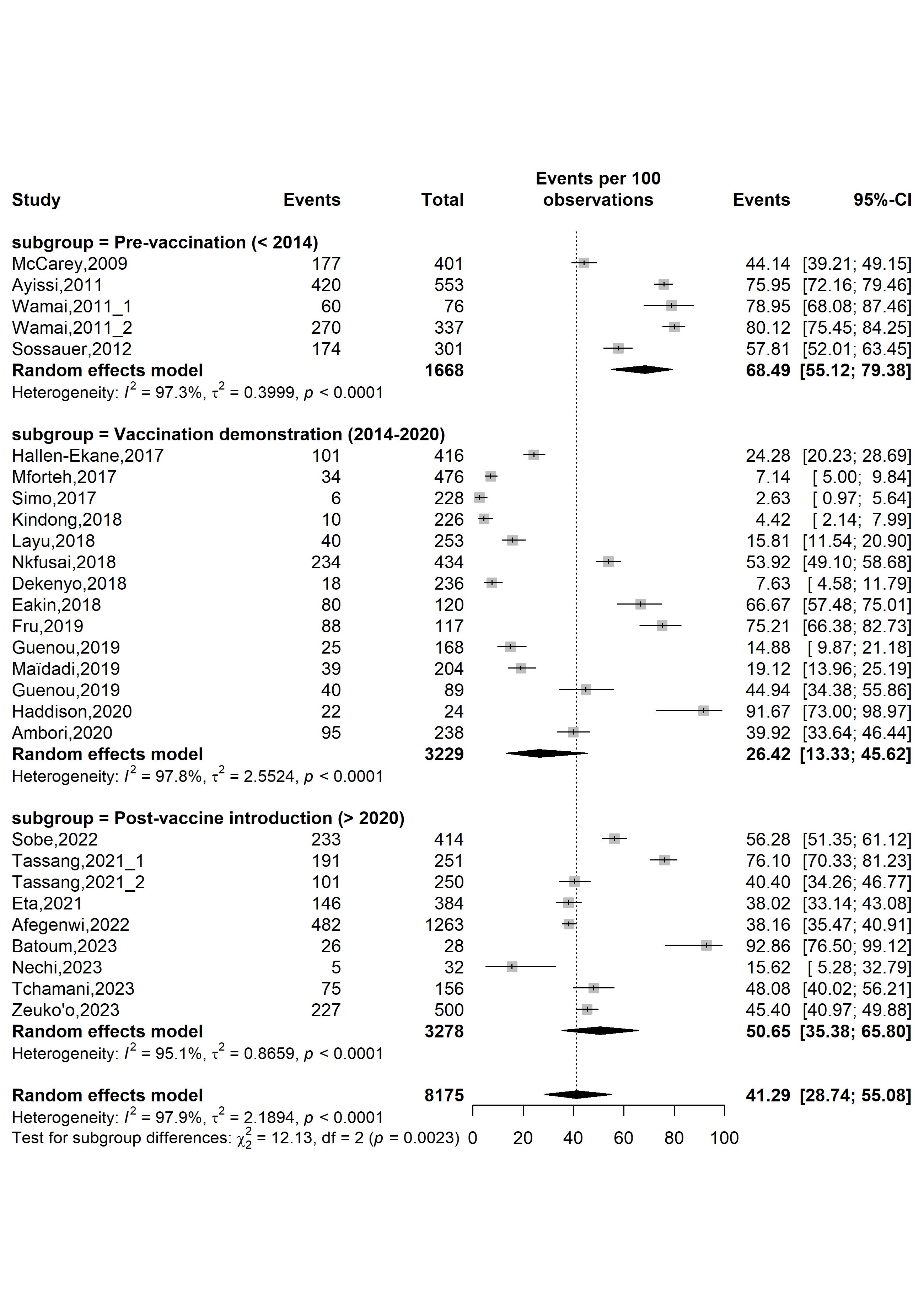

**Supplementary Fig. 6** Pooled prevalence of human papillomavirus (HPV) vaccine awareness according to specific HPV vaccine introduction timeframe in Cameroon

**Study setting**

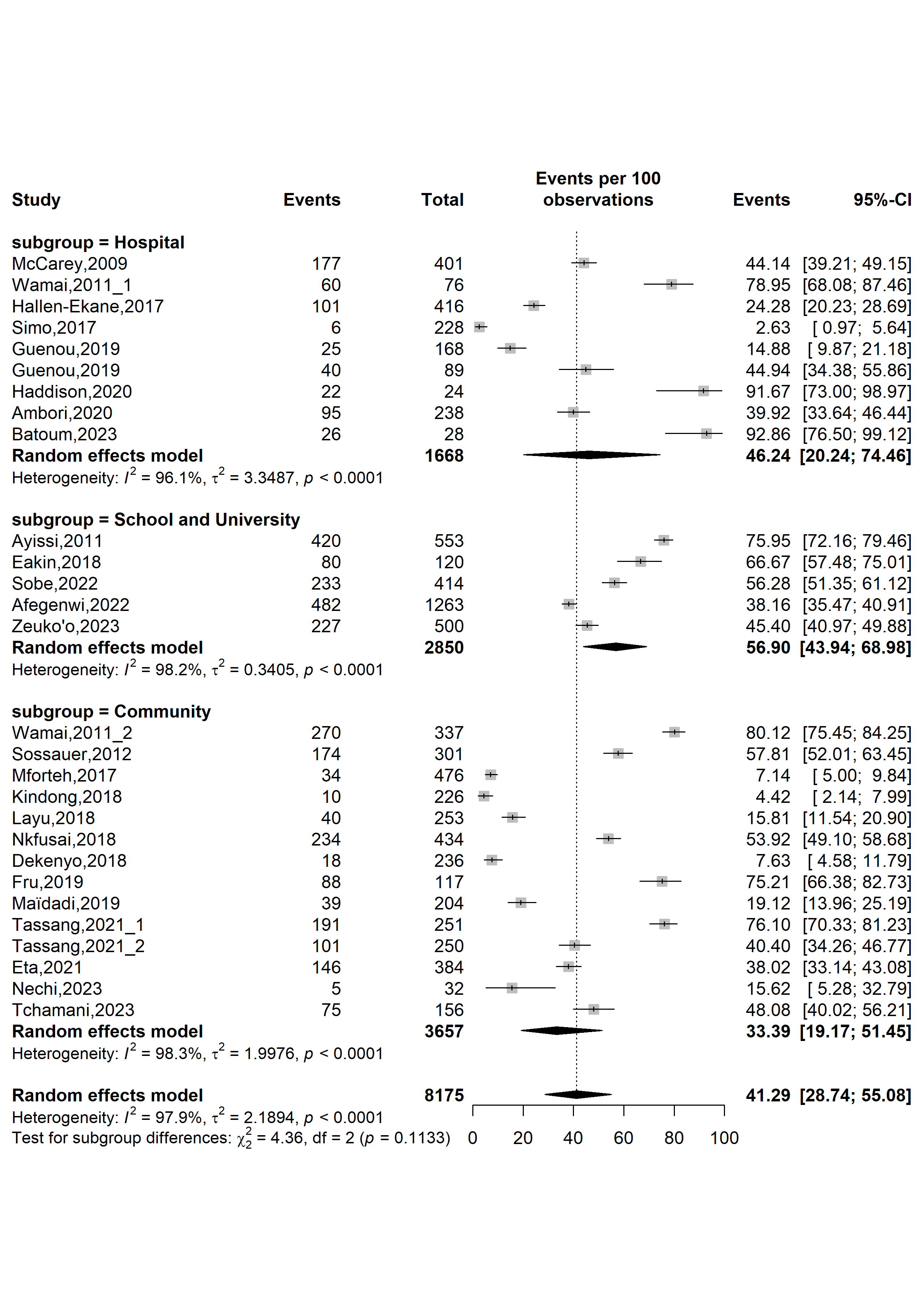

**Supplementary Fig. 7** Pooled prevalence of human papillomavirus vaccine awareness according to study settings in Cameroon

**Study site**

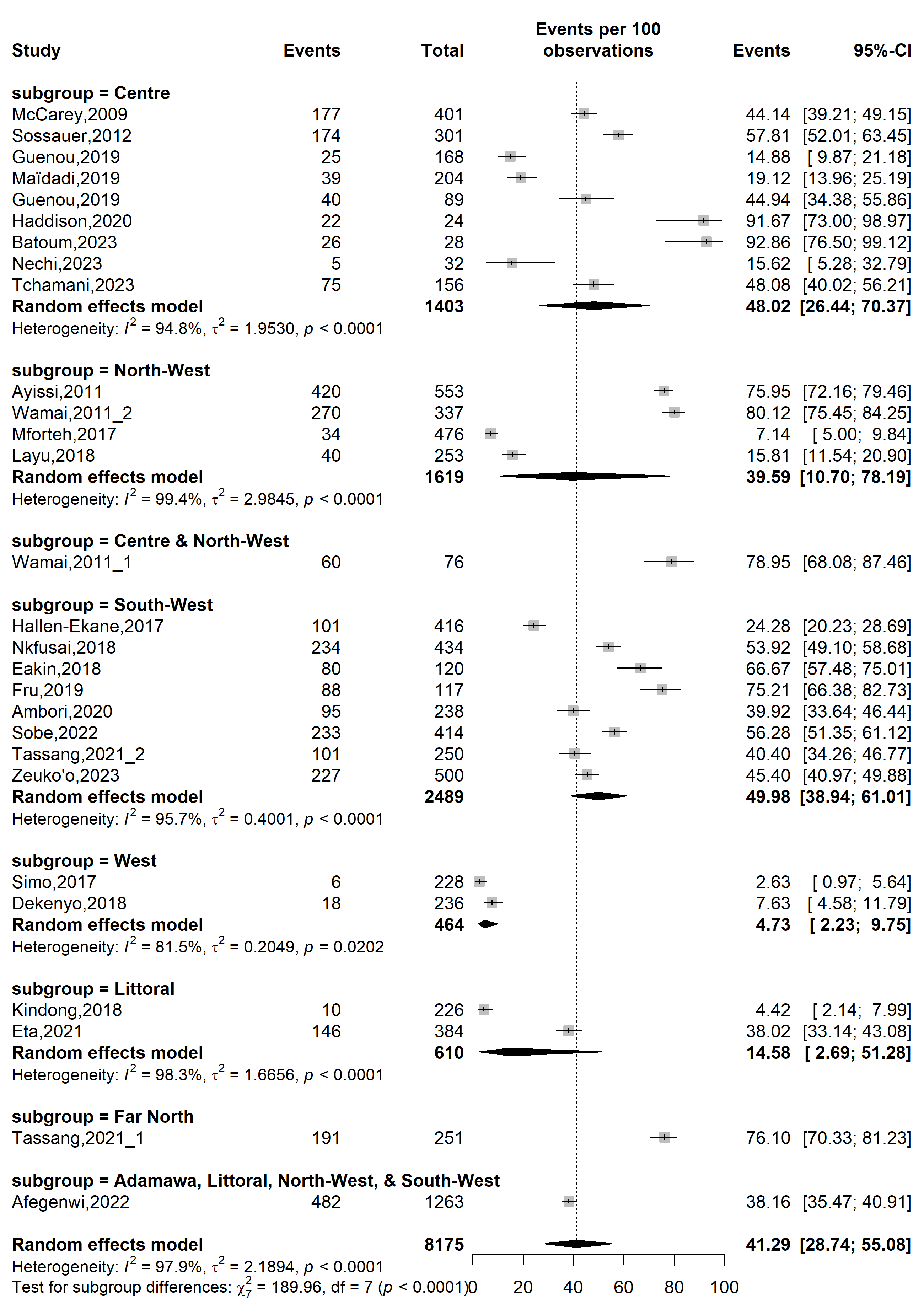

**Supplementary Fig. 8** Pooled prevalence of human papillomavirus vaccine awareness according to study sites in Cameroon

**Sampling method**

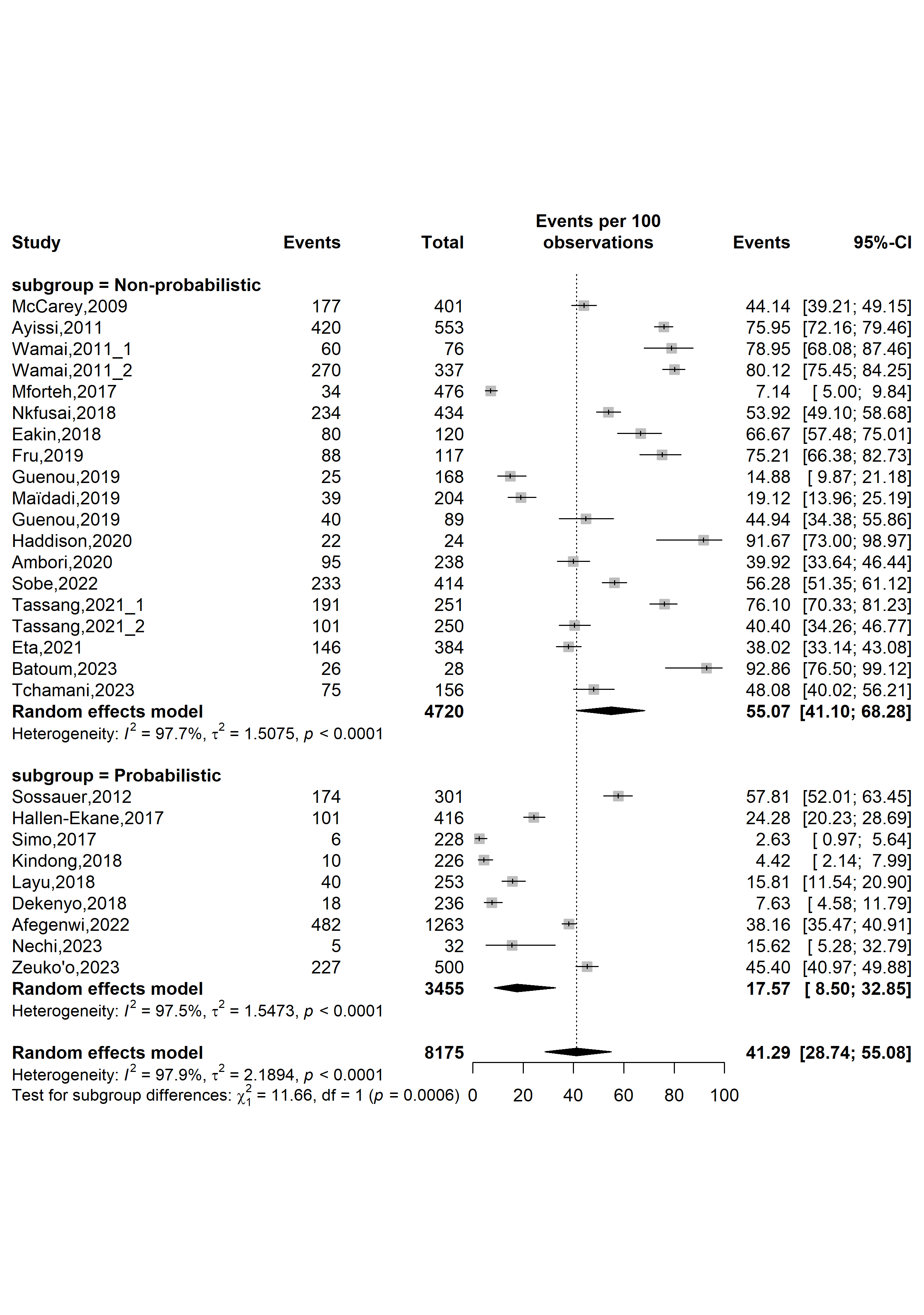

**Supplementary Fig. 9** Pooled prevalence of human papillomavirus vaccine awareness by sampling methods in Cameroon

**Type of participants**

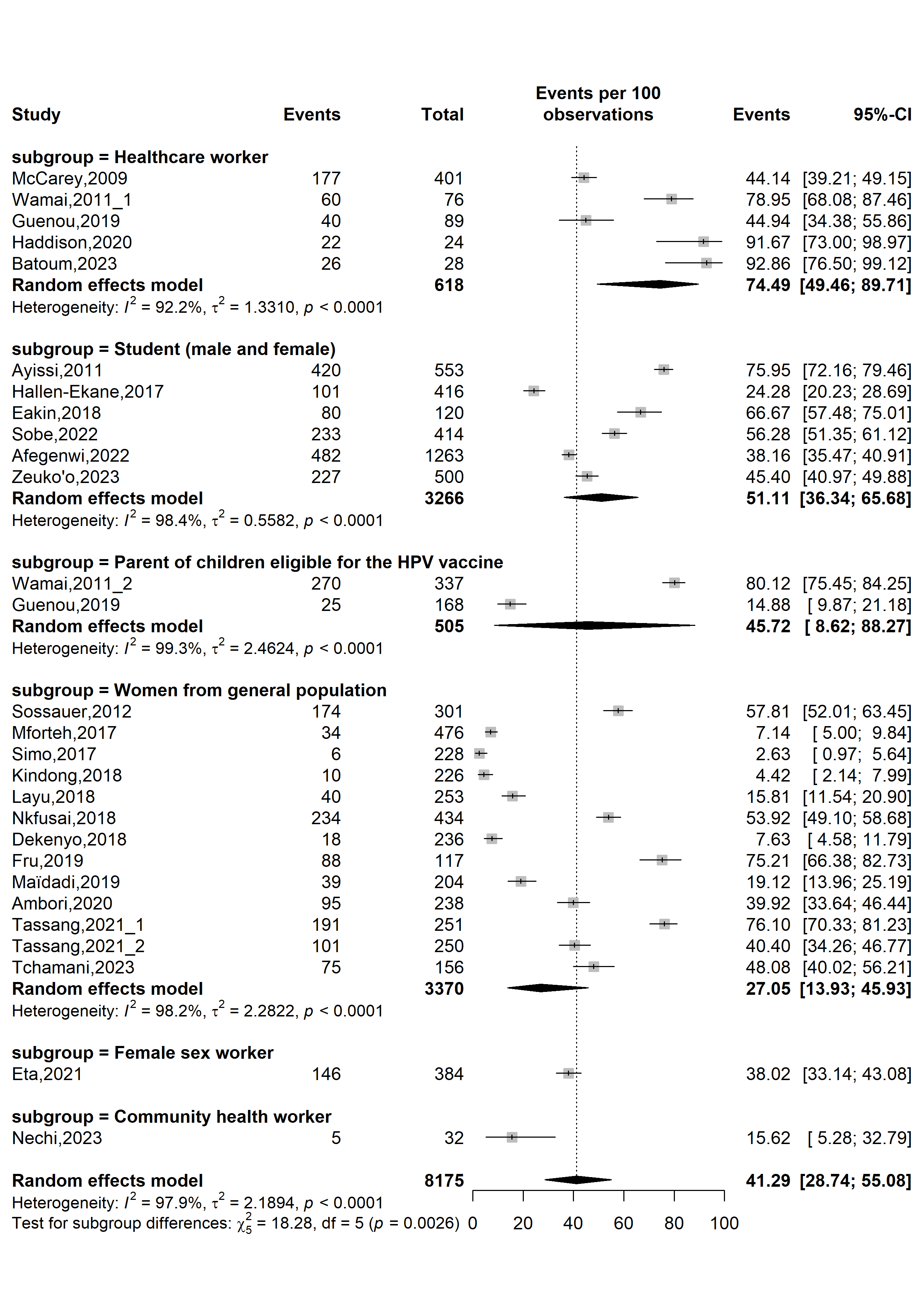

**Supplementary Fig. 10** Pooled prevalence of human papillomavirus vaccine awareness by types of participants in Cameroon

**Publication bias assessment**

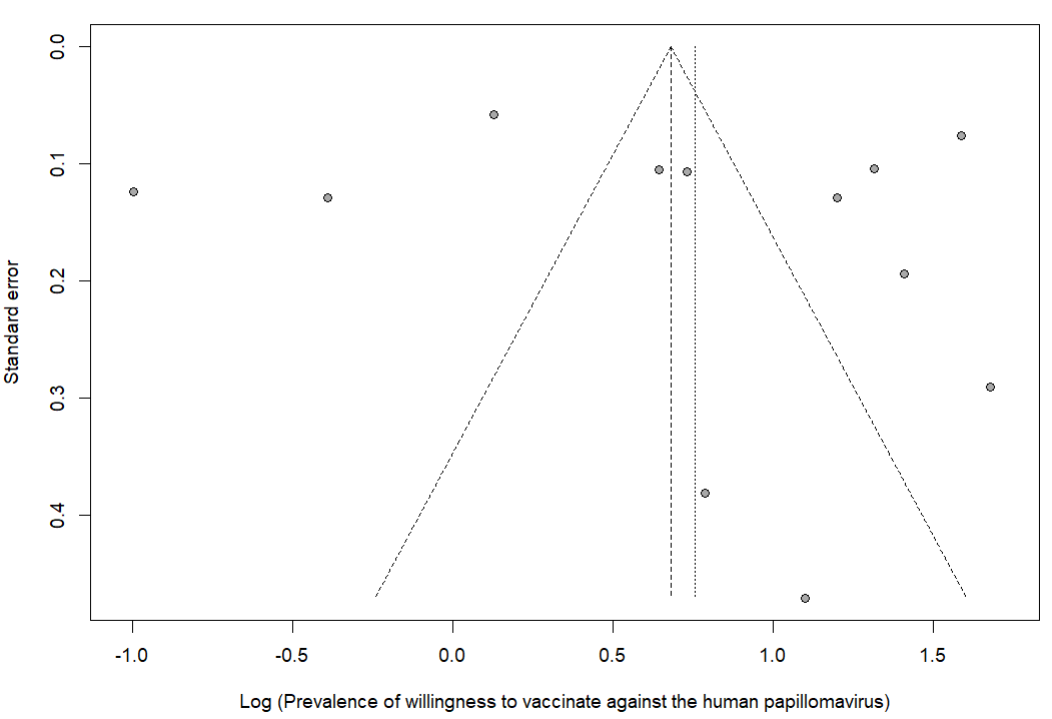

Egger’s test *p*-value = 0.707

Begg’s test *p*-value = 0.784

**Supplementary Fig. 11** Funnel plot displaying the risk of publication bias of studies assessing willingness to vaccinate against human papillomavirus in Cameroon

**Sensitivity analysis**

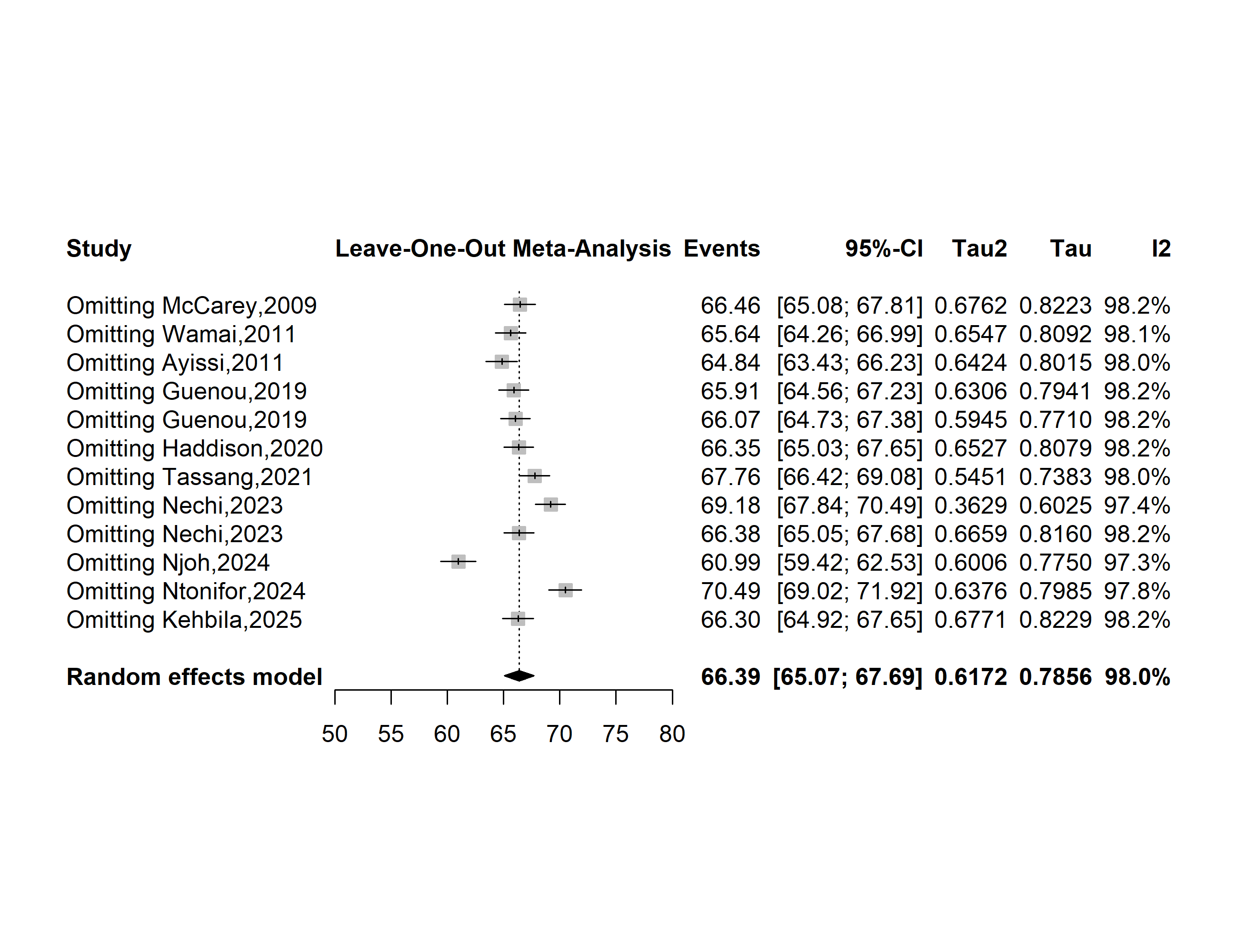

**Supplementary Fig. 12** Sensitivity analysis of the pooled prevalence of willingness to vaccinate against human papillomavirus in Cameroon

**Publication bias assessment**

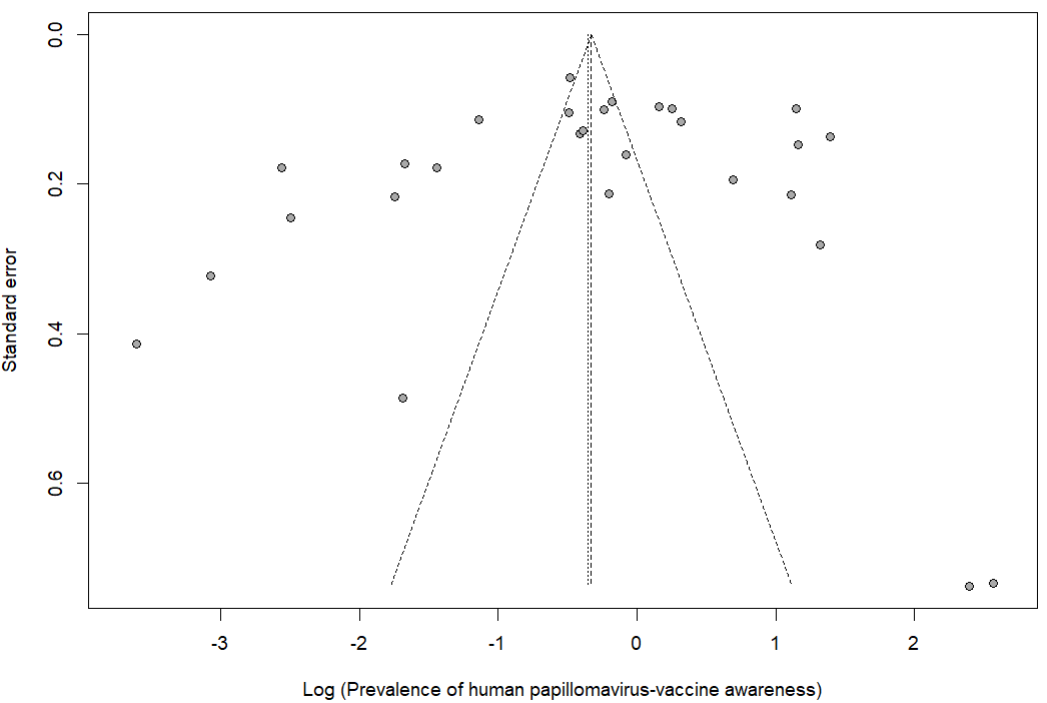

Egger’s test *p*-value = 0.455

Begg’s test *p*-value = 0.323

**Supplementary Fig. 13** Funnel plot displaying the risk of publication bias of studies assessing the prevalence of human papillomavirus vaccine awareness in Cameroon

**Sensitivity analysis**

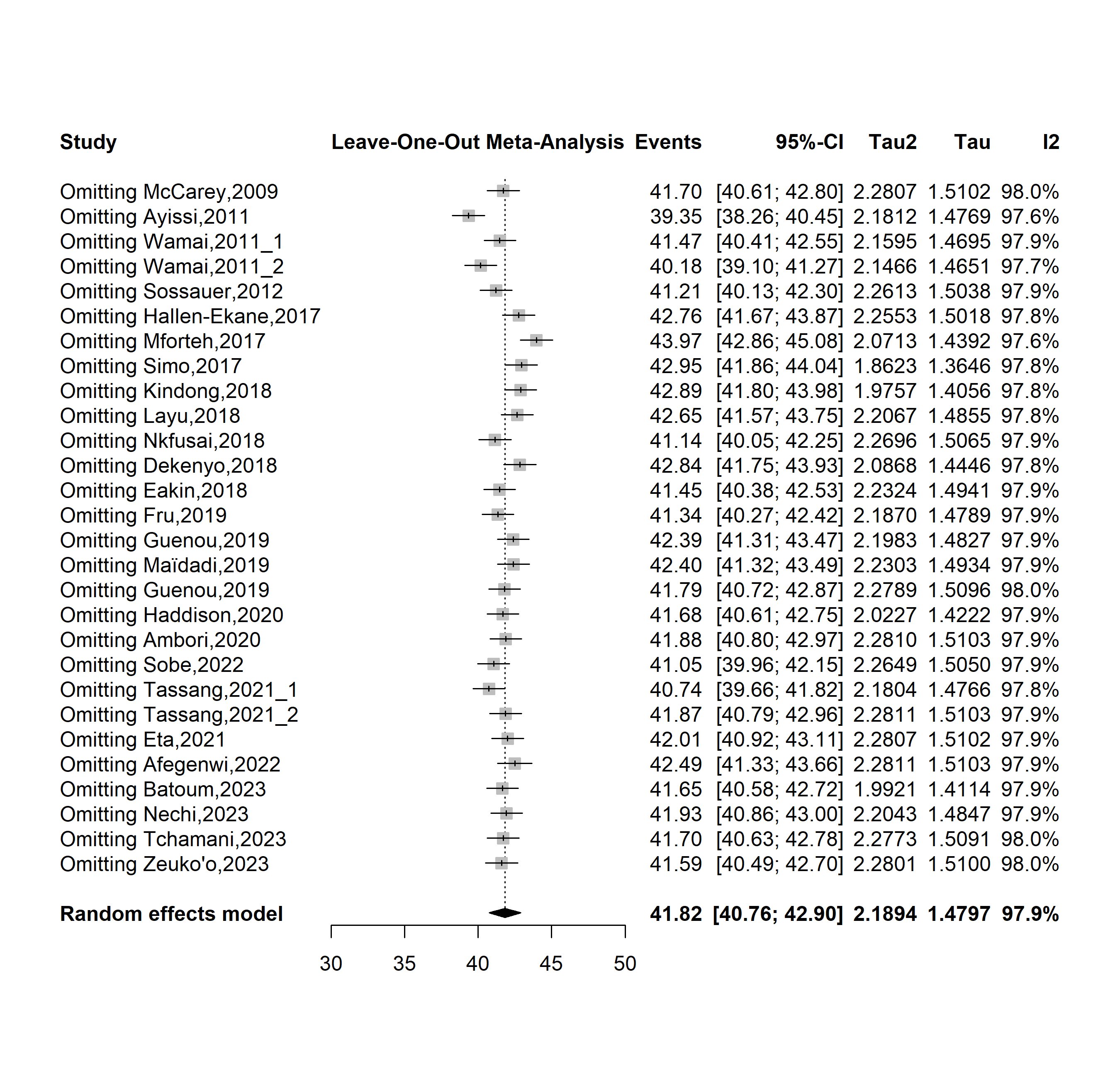

**Supplementary Fig. 14** Sensitivity analysis of the pooled prevalence of human papillomavirus vaccine awareness in Cameroon

**Sensitivity analysis**

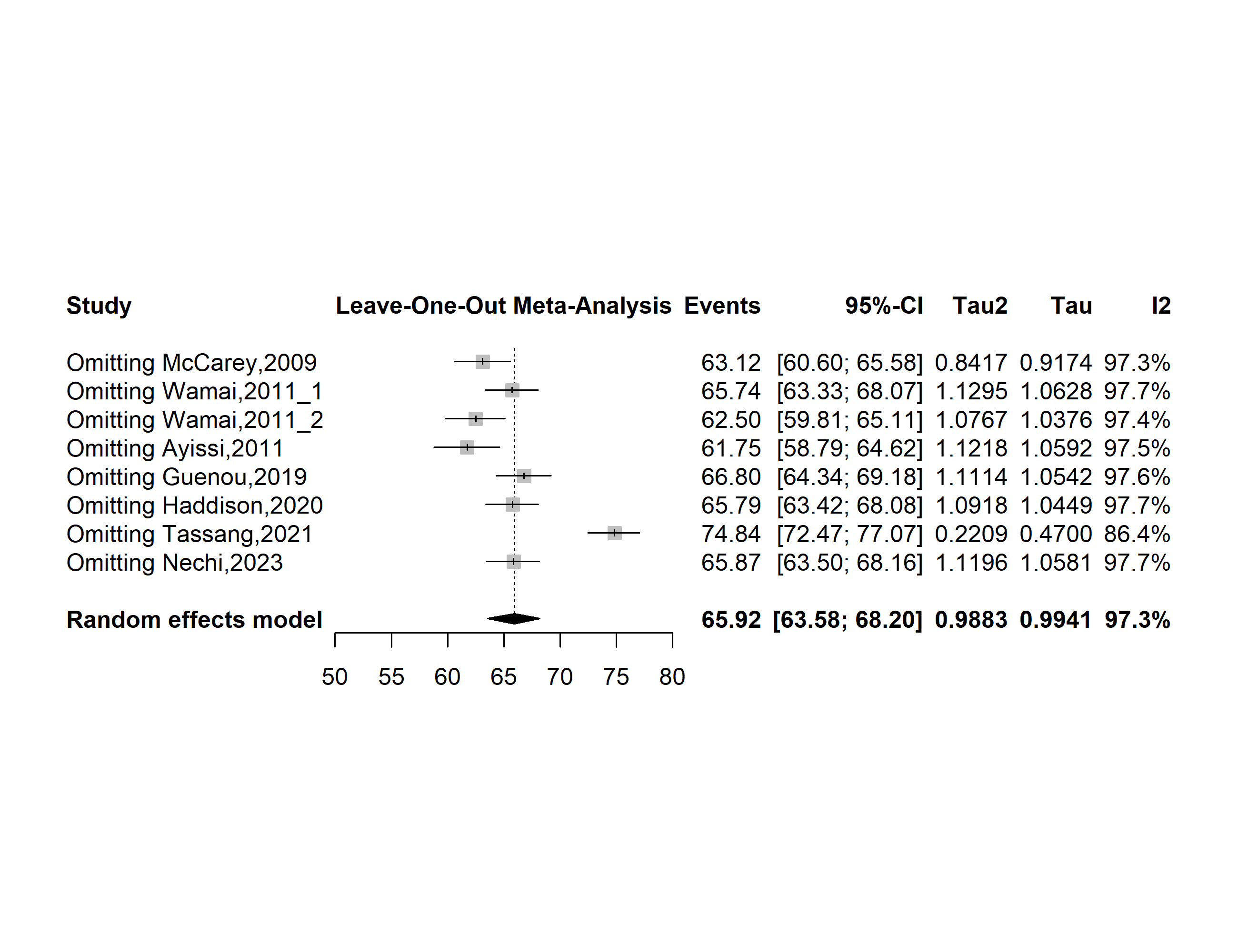

**Supplementary Fig. 15** Forest plot displaying the sensitivity analysis of pooled the prevalence of human papillomavirus vaccine recommendation in Cameroon

**Publication bias assessment**

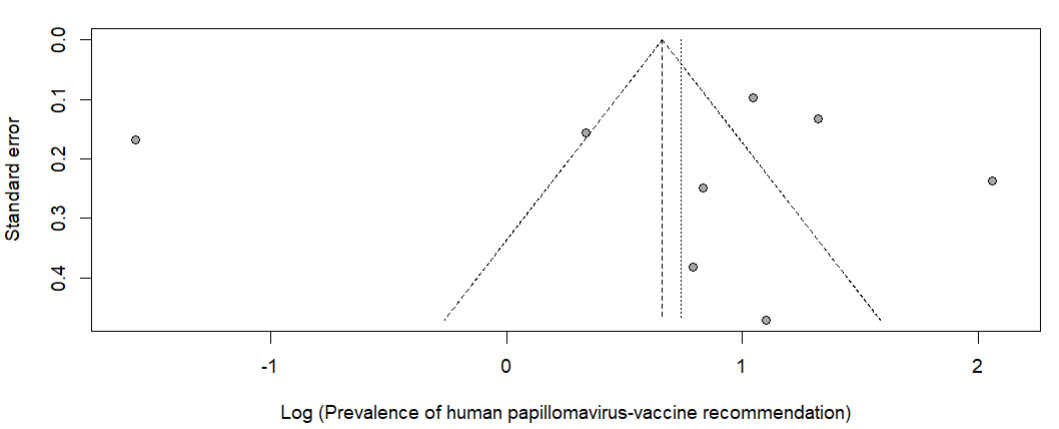

**Supplementary Fig. 16** Funnel plot displaying the risk of publication bias of studies assessing the prevalence of human papillomavirus vaccine recommendation in Cameroon

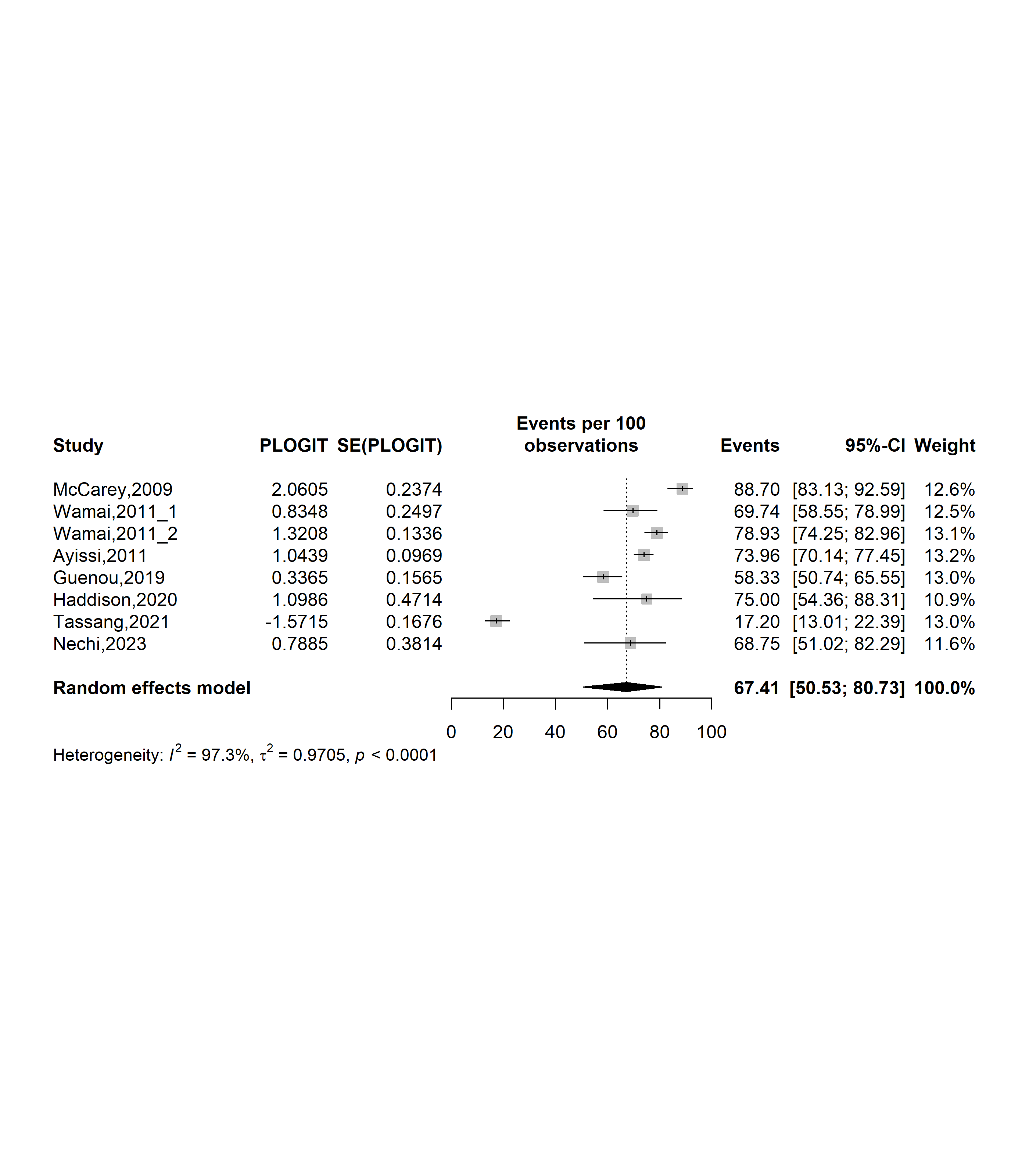

**Supplementary Fig. 17** Forest plot displaying the trim and fill analysis of studies assessing the prevalence of human papillomavirus vaccine recommendation in Cameroon

**Sensitivity analysis**

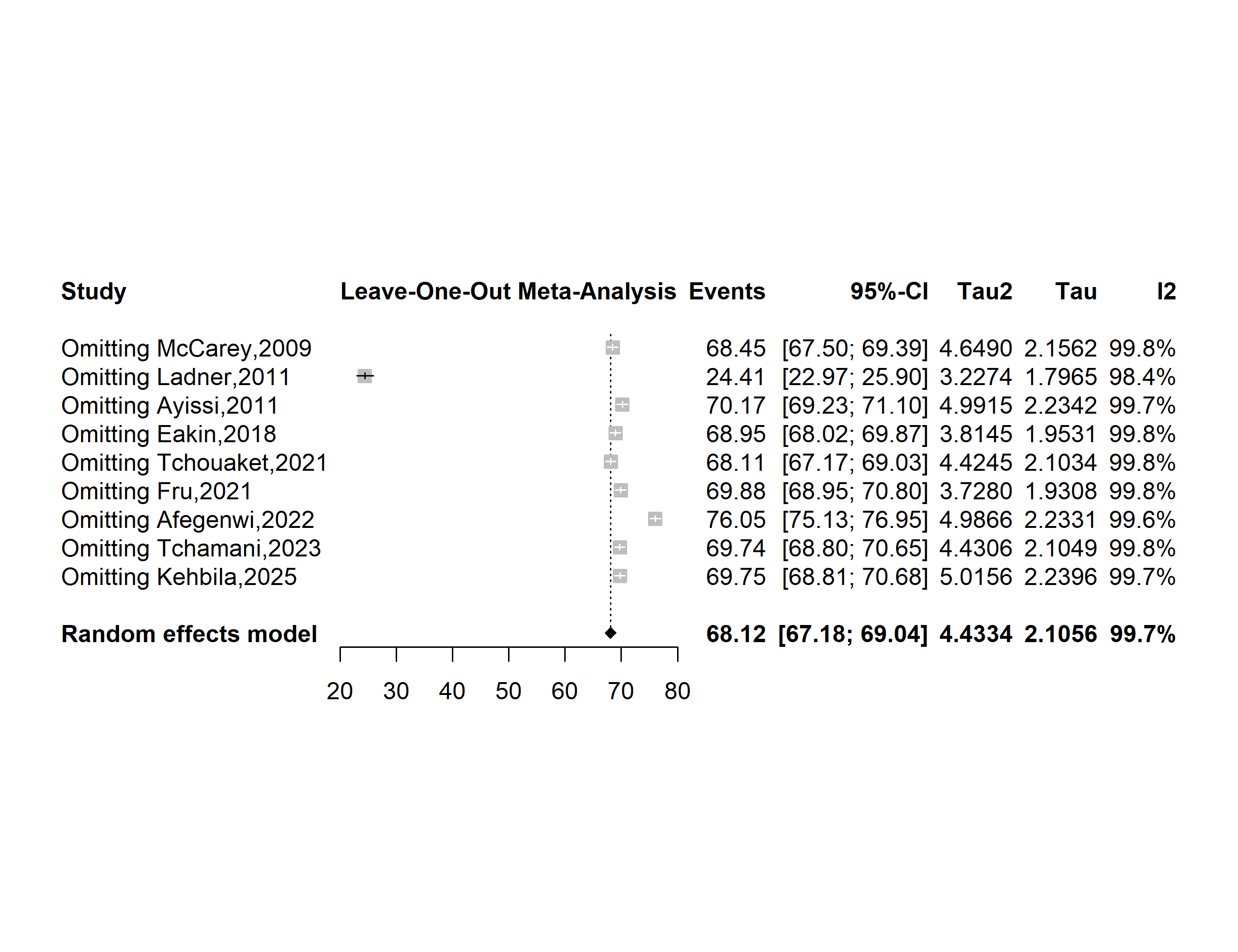

**Supplementary Fig. 18** Forest plot displaying the sensitivity analysis of pooled the prevalence of human papillomavirus vaccine uptake in Cameroon

**Publication bias assessment**

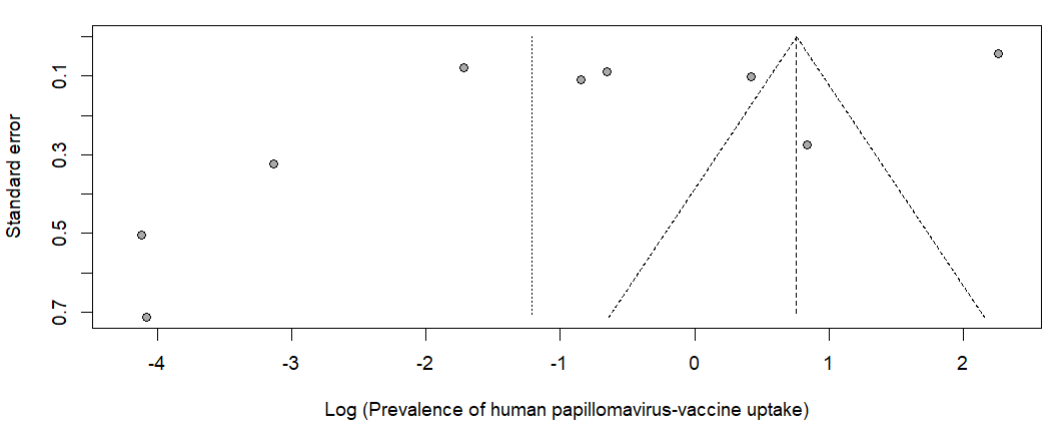

**Supplementary Fig. 19** Funnel plot displaying the risk of publication bias of studies assessing the prevalence of human papillomavirus vaccine uptake in Cameroon

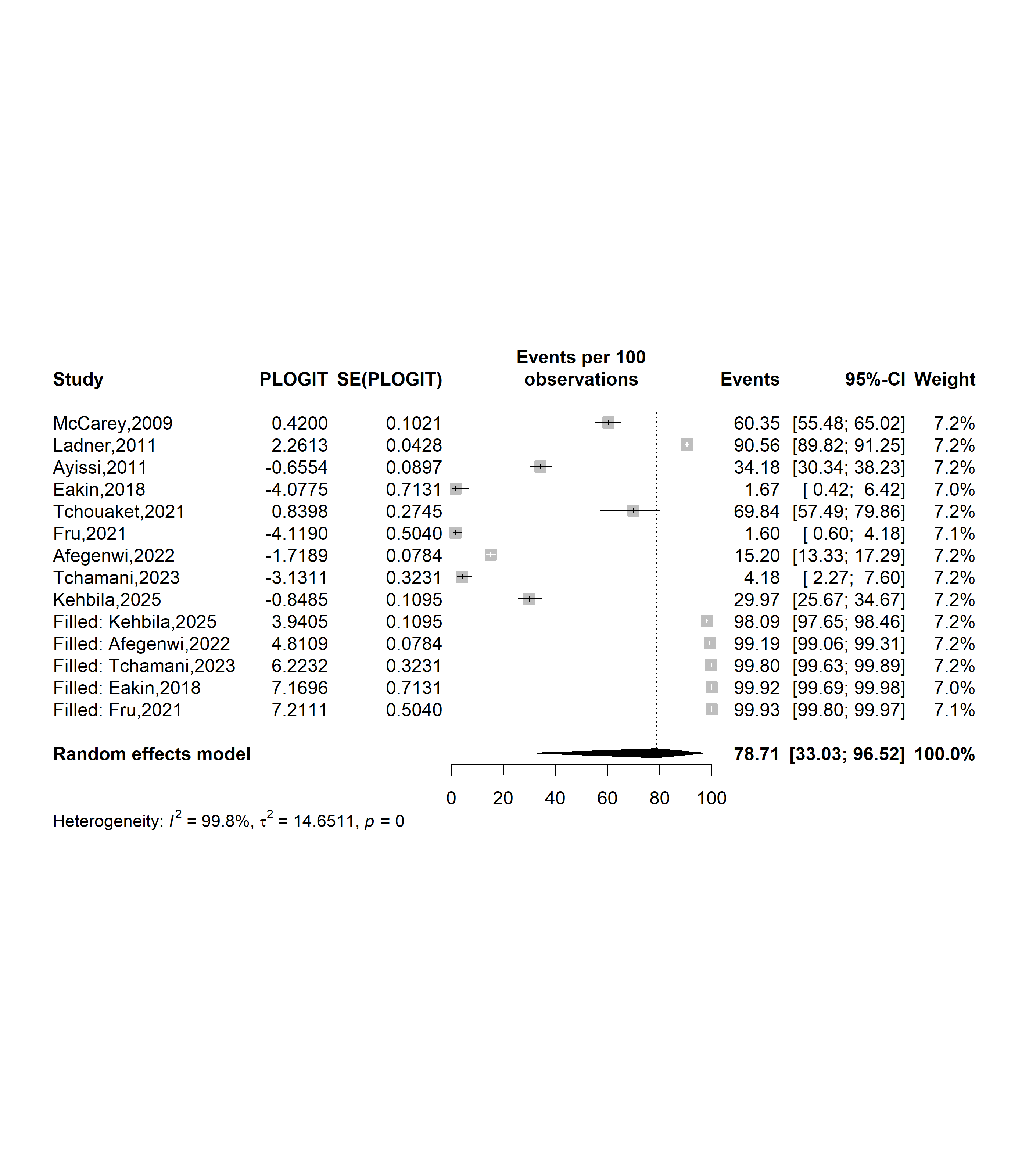

**Supplementary Fig. 20** Forest plot displaying the trim and fill analysis of studies assessing the prevalence of human papillomavirus vaccine uptake in Cameroon

**Predictor of HPV vaccine hesitancy**

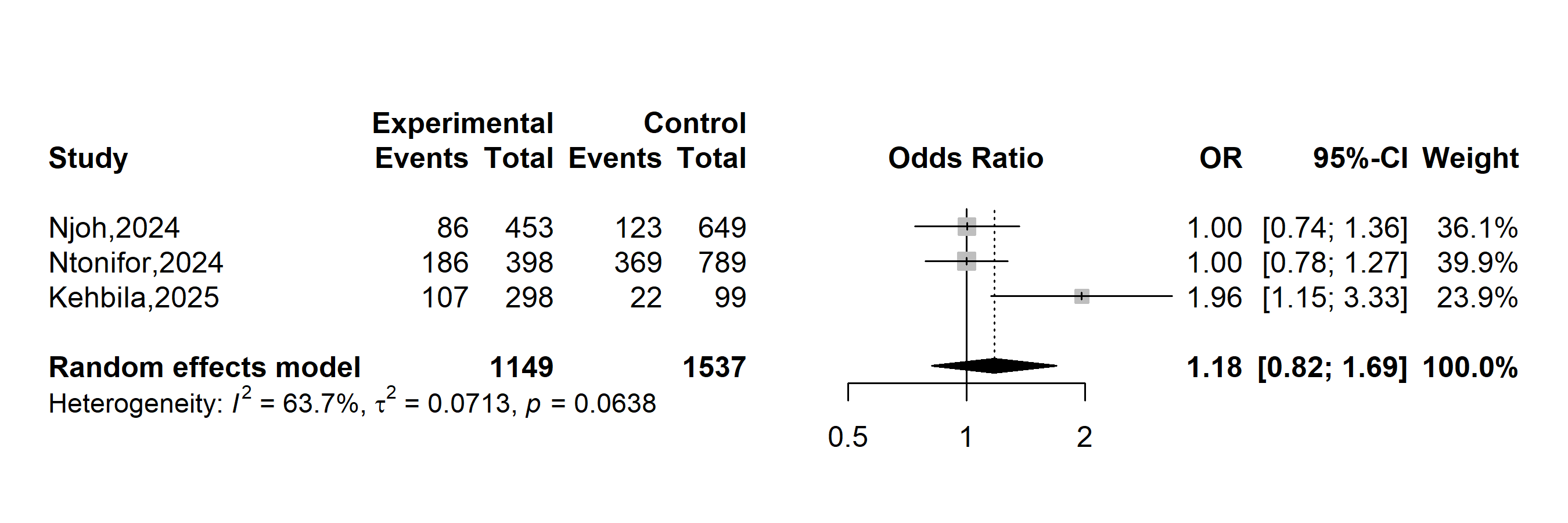

**Supplementary Fig. 21** Pooled odds ratio hesitancy to vaccinate against human papillomavirus in Cameroon (Gender: female vs. male)

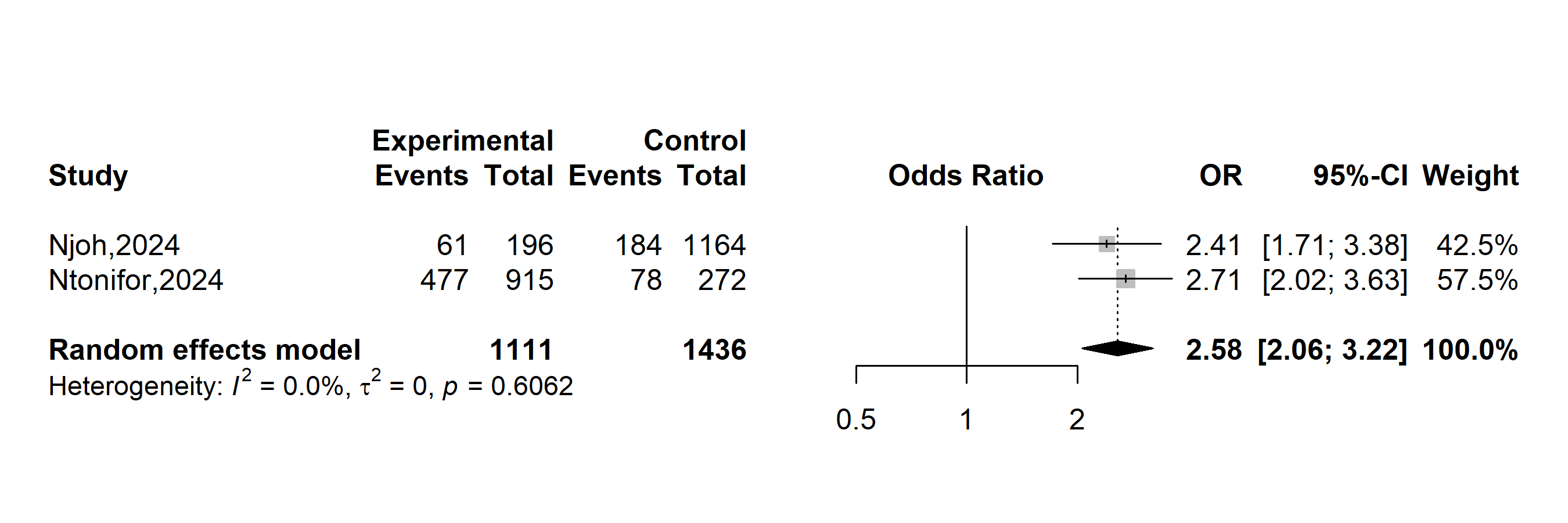

**Supplementary Fig. 22** Pooled odds ratio of hesitancy to vaccinate against human papillomavirus (HPV) in Cameroon (Individual knowledge of HPV: no vs. yes)

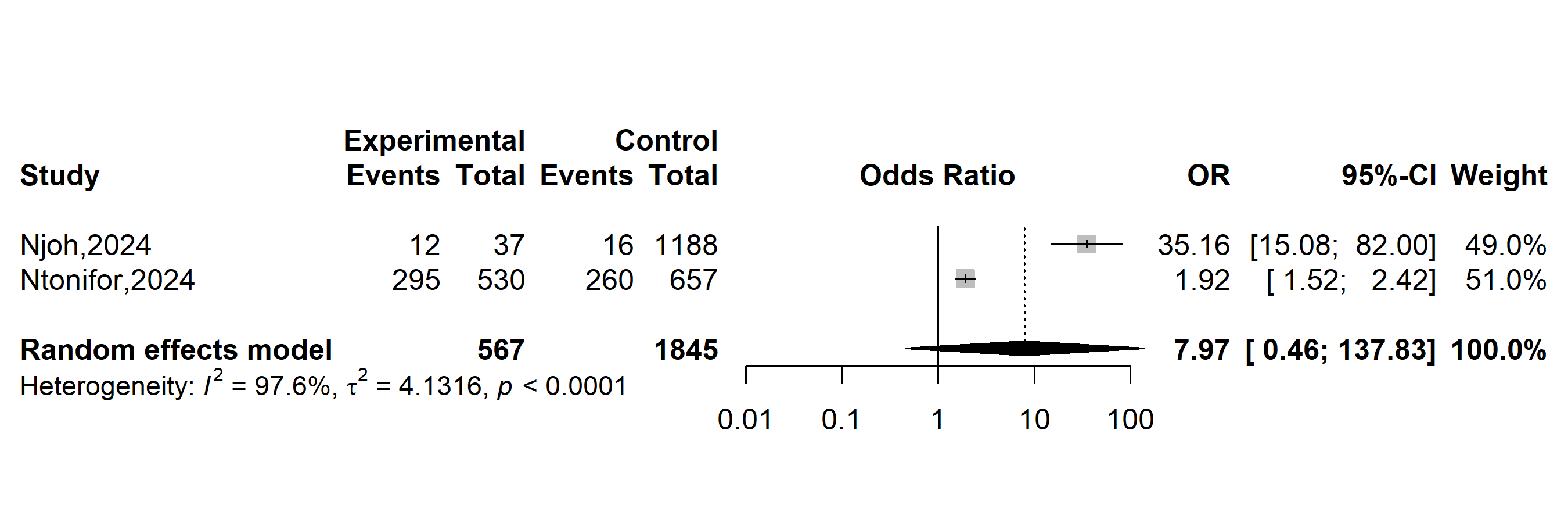

**Supplementary Fig. 23** Pooled odds ratio of hesitancy to vaccinate against human papillomavirus in Cameroon (Individual knowledge of cervical cancer: no vs. yes)
